# COVID-19 Hospitalisation in Portugal, the first year: Results from hospital discharge data

**DOI:** 10.1101/2022.03.03.22271349

**Authors:** João F. Pereira, Constantino Caetano, Liliana Antunes, Paula Patrício, Maria Luísa Morgado, Baltazar Nunes

## Abstract

Using Portuguese diagnosis-related groups (DRGs) data we analysed the dynamics of the COVID-19 hospitalisation in Portugal, with the ultimate goal of estimating parameters to be used in COVID-19 mathematical modelling.

After processing the data, corresponding to the period between March 2020 and the end of March 2021, an exploratory analysis was conducted, in which the length of stay (LoS) in hospital care, the time until death (TuD), the number of admissions and mortality count were estimated and described with respect to the age and gender of the patients, but also by region of admission and evolution over time.

The median and mean of LoS was estimated to be 8 and 12.5 days (IQR:4-15) for non-ICU patients and for ICU patients as 18 and 24.3 days (IQR:11-30). The percentage of patients that, during their hospitalisation, required ICU care was determined to be 10.9%. In-hospital mortality of non-ICU patients (22.6%) and of ICU patients (32.8%) were also calculated. In a visual exploratory analysis we observed that they changed with time, age group, gender and region.

Various univariable probability distributions were fitted to the LoS and TuD data, using maximum likelihood estimation (MLE) but also maximum goodness-of-fit estimation (MGE) to obtain the distribution parameters.

**AMS Subject Classification:** 62-07,62E07,62F10,62P10

## 1. INTRODUCTION

In Portugal, the first case of COVID-19 was reported on March 2nd, 2020. On March 12, 2020 the Portuguese government declared the State of Emergency, and lockdown measures were implemented to stop the rapid increase of infections. From then onwards, several periods of confinement were implemented, according to the rise and fall in the number of cases and other epidemiological factors [2, 6].

Mathematical models are a suitable tool to develop scenarios to evaluate the impact of different control measures. For COVID-19, several mathematical models have been developed (see [11] and the references therein). The success of these models in capturing the transmission dynamics strongly depend on a good parametrisation. While it is expected that disease and biological parameters do not differ among different regions, other, as demographic and social related parameters, strongly depend on the population under study. Furthermore, there was a necessity for models to consider the hospitalisation dynamics of COVID-19, in order to assess health care burden. To this end, we analysed Portuguese DRG (diagnosis-related group) data with two main goals: to characterise hospital admissions and to estimate suitable parameters of interest.

In Portugal, the clinical data of hospitalized patients is logged according to the International Classification of Diseases, Clinical Modification and International Classification of Diseases, Tenth Revision Procedure Coding System (ICD-10-CM/PCS), in line with the 2020/2021 normative [3].This unstructured data is then grouped into DRGs, with the objective of being used in healthcare management, pricing and decision-making by the hospital management and health professionals, but also by authorized national agencies that use it to perform various types of public health studies and statistical analysis on hospital treatment. The DRG system currently in use in Portugal is the *all-patient refined diagnosis-related groups* (hereafter APR-DRG) (*Administração Central do Sistema de Saúde* 2014). This system assigns codes to the patient which refers to a specific diagnosis, so that patients with similar resource usage and costs are clustered. According to Julio Souza *et.al* [19], the overall credibility of the Portuguese data is very high and the data can be considered credible and usable, despite some particular cases of diagnosis that have less cohesive date.

This paper is organized as follows: In Section 2 the original DRG data and the transformed data used in this study are explained as well as the description of the methodology used in the exploratory analysis and in the fitting of the distributions. In Section 3 the exploratory analysis is performed, in which the LoS and TuD, number of entries, fatalities and other parameters, for either ICU and non-ICU patients, is characterized and compared between genders, age groups and regions, as well as their evolution over time. In Section 4 the results from this study are discussed. An appendix section A is also provided, with an extended explanation of the data and all transformations carried out and also more extensive graphs and tables and other relevant information.

## 2. METHODS

### 2.1. Data

The data used in this study was obtained from the BI-MH platform of the SPMS/ACSS (Ministry of Health Shared Services / Central Administration of the Health System, IP) services of Portugal and is composed by the DRG data, with hospital discharges, having a COVID-19 diagnosis.

The DRG data was restructured in order to obtain the age of the patients and their respective age group, the regional health administration (ARS) to which the hospital belongs, LoS and TuD, as explained in the appendix A.1. This new data, containing 54685 entries, is described in appendix A.2, where the used variables are identified and explained. Because the DRG data is recorded only when a patient is discharged, a right truncation problem arises. In order to overcome this, we disregard admissions from the last two months, as explained in the appendix A.3. As such the considered data ranges are from 1 of March 2020 to 31 of March 2021. Additionally, from this restructured data, the admissions were aggregated by day. The type of hospitalisation is distinguished between:

- **Non-ICU patients:** patients who have not received treatment in intensive care units during their stay in the hospital;
- **ICU patients:** patients who received treatment intensive care units during their hospitalisation.

The regions used follow the areas of the regional health administrations (ARSs) established by the Portuguese national health service (SNS). The population numbers for each region were extracted from the Portuguese Central Administration of the Health Service (ACSS) website (data from June 2016 link1). The five considered ARSs that cover Continental Portugal are: ARS Norte, ARS Centro, ARS Lisboa e Vale do Tejo (LVT), ARS Alentejo and ARS Algarve. The Portuguese autonomous regions (Açores and Madeira) are not included in the study due to lack of data.

The population estimates (total and by gender) correspond to the resident population in Continental Portugal and were extracted from the census 2021 data, elaborated by *Statistics Portugal* (INE), the Portuguese official statistical institution, and is avaliable at INE website at link2. The population numbers by age group correspond to the provisional resident population data obtained from the census data, available at the link3.

A table containing the number of hospital entries, the population number for Portugal and percentages calculated from these values was used in several analysis, being presented in the appendix A.1 in table 3. The daily data is presented in appendix A.2 tables 4,5,6,7 and 8.

**Table 1:** Variable identification of the provided DRG data

| Variable | Description |
| --- | --- |
| Health institution | Institution where the entries took place |
| Gender | Gender of the patient |
| Date of birth | Birth date of the patient |
| Hospital process | Anonymised data related to the hospital process number |
| Hospital episode | Anonymised data related to the hospital episode number |
| Destination after discharge | Destination of the patient after medical discharge, including death. |
| Discharge date | Date the patient was discharged from the hospital |
| Entry date | Date the patient entered hospital care |
| ICU | Whether or not the patient was in ICU care |
| Type of episode | Time of stay in hospital care groups (short term, normal, long term and prolonged evolution) |
| Diagnostic code | Diagnosis-related groups (DRG) code, in this case all entries have the U071 code, that identifies COVID-19 diagnosis. |
| Diagnostic | Diagnostic represented by the Diagnostic code, in this case all entries have the COVID-19 diagnostic |
| Diagnostic order | Order of diagnostic of the patient in that day |

**Table 2:** Variable identification of the used data, restructured from the DRG data

| Variable | Description |
| --- | --- |
| Region | ARS of the health institution (Norte, Centro, LVT, Alentejo e Algarve) |
| Age | Age of the patient, at the time of entry |
| Gender | Gender of the patient |
| Entry date | Patient entry date to the health institution |
| Discharge date | Patient discharge date from the health institution |
| Destination after discharge | Destination after medical discharge, includes as a outcome death. |
| Length of stay | Length of stay in health care, corresponds to number of days between the discharge date and the entry date, in days |
| Group | Age group corresponding to the age of the patient |

**Table 3:**
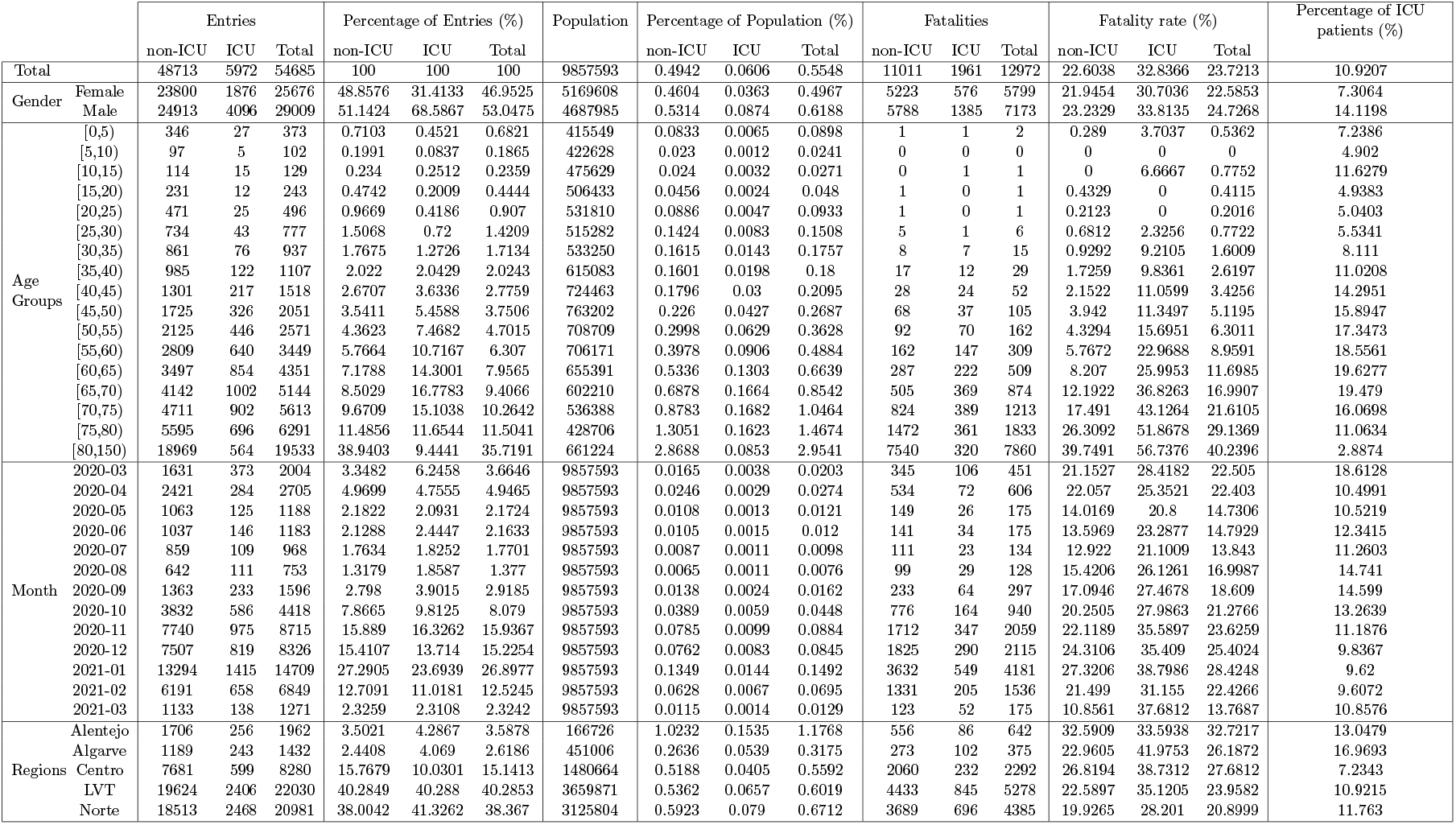
Table with various data in relation to gender, age group, time and region.

**Table 4:**
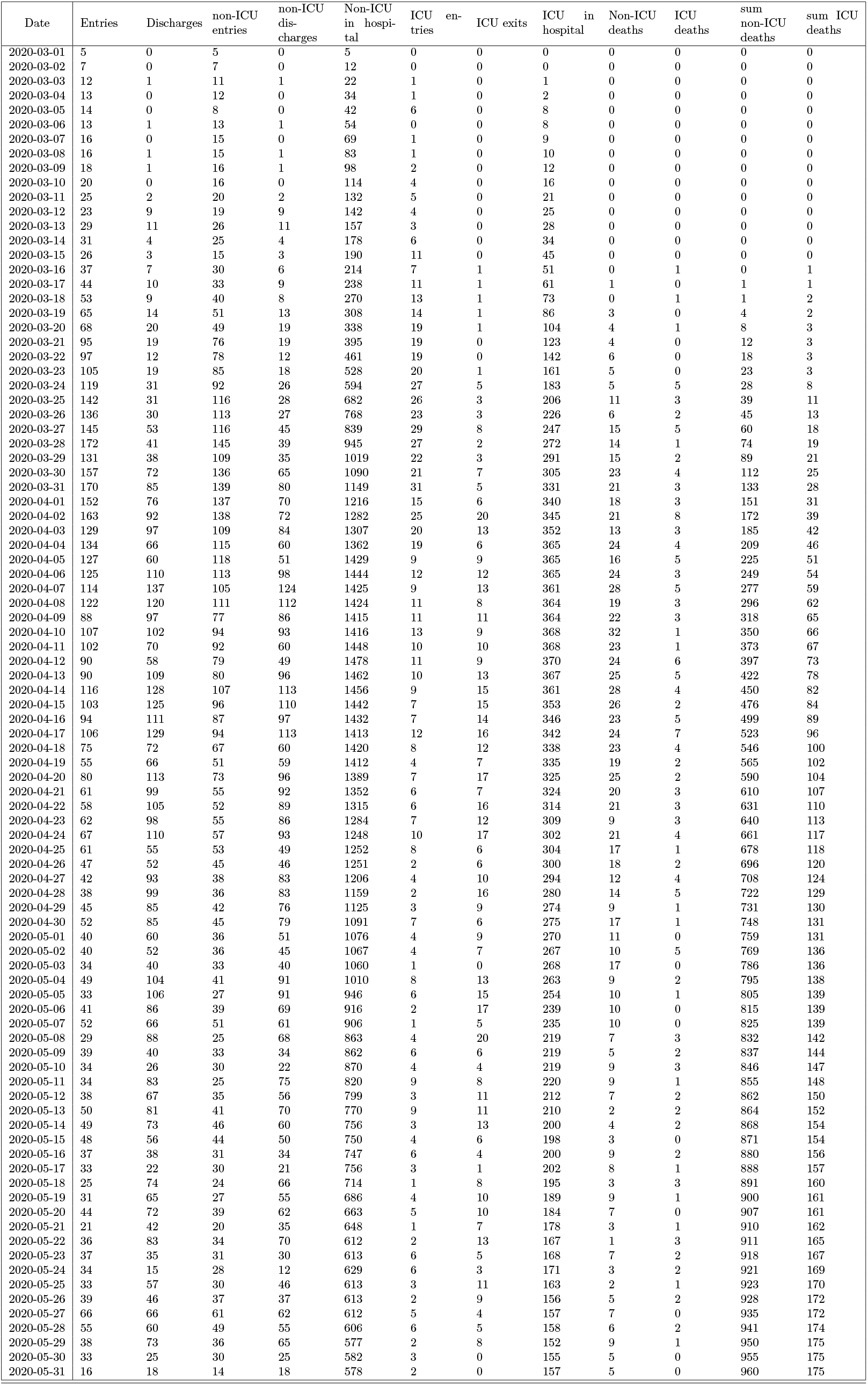
Table with daily admissions (Part I)

| Date | Entries | Discharges | non-ICU<br>entries | non-ICU<br>dis-<br>charges | Non-ICU<br>in hospi-<br>tal | ICU en-<br>tries | ICU exits | ICU in<br>hospital | Non-ICU<br>deaths | ICU<br>deaths | sum<br>non-ICU<br>deaths | sum ICU<br>deaths |
| --- | --- | --- | --- | --- | --- | --- | --- | --- | --- | --- | --- | --- |
| 2020-03-01 | 5 | 0 | 5 | 0 | 5 | 0 | 0 | 0 | 0 | 0 | 0 | 0 |
| 2020-03-02 | 7 | 0 | 7 | 0 | 12 | 0 | 0 | 0 | 0 | 0 | 0 | 0 |
| 2020-03-03 | 12 | 1 | 11 | 1 | 22 | 1 | 0 | 1 | 0 | 0 | 0 | 0 |
| 2020-03-04 | 13 | 0 | 12 | 0 | 34 | 1 | 0 | 2 | 0 | 0 | 0 | 0 |
| 2020-03-05 | 14 | 0 | 8 | 0 | 42 | 6 | 0 | 8 | 0 | 0 | 0 | 0 |
| 2020-03-06 | 13 | 1 | 13 | 1 | 54 | 0 | 0 | 8 | 0 | 0 | 0 | 0 |
| 2020-03-07 | 16 | 0 | 15 | 0 | 69 | 1 | 0 | 9 | 0 | 0 | 0 | 0 |
| 2020-03-08 | 16 | 1 | 15 | 1 | 83 | 1 | 0 | 10 | 0 | 0 | 0 | 0 |
| 2020-03-09 | 18 | 1 | 16 | 1 | 98 | 2 | 0 | 12 | 0 | 0 | 0 | 0 |
| 2020-03-10 | 20 | 0 | 16 | 0 | 114 | 4 | 0 | 16 | 0 | 0 | 0 | 0 |
| 2020-03-11 | 25 | 2 | 20 | 2 | 132 | 5 | 0 | 21 | 0 | 0 | 0 | 0 |
| 2020-03-12 | 23 | 9 | 19 | 9 | 142 | 4 | 0 | 25 | 0 | 0 | 0 | 0 |
| 2020-03-13 | 29 | 11 | 26 | 11 | 157 | 3 | 0 | 28 | 0 | 0 | 0 | 0 |
| 2020-03-14 | 31 | 4 | 25 | 4 | 178 | 6 | 0 | 34 | 0 | 0 | 0 | 0 |
| 2020-03-15 | 26 | 3 | 15 | 3 | 190 | 11 | 0 | 45 | 0 | 0 | 0 | 0 |
| 2020-03-16 | 37 | 7 | 30 | 6 | 214 | 7 | 1 | 51 | 0 | 1 | 0 | 1 |
| 2020-03-17 | 44 | 10 | 33 | 9 | 238 | 11 | 1 | 61 | 1 | 0 | 1 | 1 |
| 2020-03-18 | 53 | 9 | 40 | 8 | 270 | 13 | 1 | 73 | 0 | 1 | 1 | 2 |
| 2020-03-19 | 65 | 14 | 51 | 13 | 308 | 14 | 1 | 86 | 3 | 0 | 4 | 2 |
| 2020-03-20 | 68 | 20 | 49 | 19 | 338 | 19 | 1 | 104 | 4 | 1 | 8 | 3 |
| 2020-03-21 | 95 | 19 | 76 | 19 | 395 | 19 | 0 | 123 | 4 | 0 | 12 | 3 |
| 2020-03-22 | 97 | 12 | 78 | 12 | 461 | 19 | 0 | 142 | 6 | 0 | 18 | 3 |
| 2020-03-23 | 105 | 19 | 85 | 18 | 528 | 20 | 1 | 161 | 5 | 0 | 23 | 3 |
| 2020-03-24 | 119 | 31 | 92 | 26 | 594 | 27 | 5 | 183 | 5 | 5 | 28 | 8 |
| 2020-03-25 | 142 | 31 | 116 | 28 | 682 | 26 | 3 | 206 | 11 | 3 | 39 | 11 |
| 2020-03-26 | 136 | 30 | 113 | 27 | 768 | 23 | 3 | 226 | 6 | 2 | 45 | 13 |
| 2020-03-27 | 145 | 53 | 116 | 45 | 839 | 29 | 8 | 247 | 15 | 5 | 60 | 18 |
| 2020-03-28 | 172 | 41 | 145 | 39 | 945 | 27 | 2 | 272 | 14 | 1 | 74 | 19 |
| 2020-03-29 | 131 | 38 | 109 | 35 | 1019 | 22 | 3 | 291 | 15 | 2 | 89 | 21 |
| 2020-03-30 | 157 | 72 | 136 | 65 | 1090 | 21 | 7 | 305 | 23 | 4 | 112 | 25 |
| 2020-03-31 | 170 | 85 | 139 | 80 | 1149 | 31 | 5 | 331 | 21 | 3 | 133 | 28 |
| 2020-04-01 | 152 | 76 | 137 | 70 | 1216 | 15 | 6 | 340 | 18 | 3 | 151 | 31 |
| 2020-04-02 | 163 | 92 | 138 | 72 | 1282 | 25 | 20 | 345 | 21 | 8 | 172 | 39 |
| 2020-04-03 | 129 | 97 | 109 | 84 | 1307 | 20 | 13 | 352 | 13 | 3 | 185 | 42 |
| 2020-04-04 | 134 | 66 | 115 | 60 | 1362 | 19 | 6 | 365 | 24 | 4 | 209 | 46 |
| 2020-04-05 | 127 | 60 | 118 | 51 | 1429 | 9 | 9 | 365 | 16 | 5 | 225 | 51 |
| 2020-04-06 | 125 | 110 | 113 | 98 | 1444 | 12 | 12 | 365 | 24 | 3 | 249 | 54 |
| 2020-04-07 | 114 | 137 | 105 | 124 | 1425 | 9 | 13 | 361 | 28 | 5 | 277 | 59 |
| 2020-04-08 | 122 | 120 | 111 | 112 | 1424 | 11 | 8 | 364 | 19 | 3 | 296 | 62 |
| 2020-04-09 | 88 | 97 | 77 | 86 | 1415 | 11 | 11 | 364 | 22 | 3 | 318 | 65 |
| 2020-04-10 | 107 | 102 | 94 | 93 | 1416 | 13 | 9 | 368 | 32 | 1 | 350 | 66 |
| 2020-04-11 | 102 | 70 | 92 | 60 | 1448 | 10 | 10 | 368 | 23 | 1 | 373 | 67 |
| 2020-04-12 | 90 | 58 | 79 | 49 | 1478 | 11 | 9 | 370 | 24 | 6 | 397 | 73 |
| 2020-04-13 | 90 | 109 | 80 | 96 | 1462 | 10 | 13 | 367 | 25 | 5 | 422 | 78 |
| 2020-04-14 | 116 | 128 | 107 | 113 | 1456 | 9 | 15 | 361 | 28 | 4 | 450 | 82 |
| 2020-04-15 | 103 | 125 | 96 | 110 | 1442 | 7 | 15 | 353 | 26 | 2 | 476 | 84 |
| 2020-04-16 | 94 | 111 | 87 | 97 | 1432 | 7 | 14 | 346 | 23 | 5 | 499 | 89 |
| 2020-04-17 | 106 | 129 | 94 | 113 | 1413 | 12 | 16 | 342 | 24 | 7 | 523 | 96 |
| 2020-04-18 | 75 | 72 | 67 | 60 | 1420 | 8 | 12 | 338 | 23 | 4 | 546 | 100 |
| 2020-04-19 | 55 | 66 | 51 | 59 | 1412 | 4 | 7 | 335 | 19 | 2 | 565 | 102 |
| 2020-04-20 | 80 | 113 | 73 | 96 | 1389 | 7 | 17 | 325 | 25 | 2 | 590 | 104 |
| 2020-04-21 | 61 | 99 | 55 | 92 | 1352 | 6 | 7 | 324 | 20 | 3 | 610 | 107 |
| 2020-04-22 | 58 | 105 | 52 | 89 | 1315 | 6 | 16 | 314 | 21 | 3 | 631 | 110 |
| 2020-04-23 | 62 | 98 | 55 | 86 | 1284 | 7 | 12 | 309 | 9 | 3 | 640 | 113 |
| 2020-04-24 | 67 | 110 | 57 | 93 | 1248 | 10 | 17 | 302 | 21 | 4 | 661 | 117 |
| 2020-04-25 | 61 | 55 | 53 | 49 | 1252 | 8 | 6 | 304 | 17 | 1 | 678 | 118 |
| 2020-04-26 | 47 | 52 | 45 | 46 | 1251 | 2 | 6 | 300 | 18 | 2 | 696 | 120 |
| 2020-04-27 | 42 | 93 | 38 | 83 | 1206 | 4 | 10 | 294 | 12 | 4 | 708 | 124 |
| 2020-04-28 | 38 | 99 | 36 | 83 | 1159 | 2 | 16 | 280 | 14 | 5 | 722 | 129 |
| 2020-04-29 | 45 | 85 | 42 | 76 | 1125 | 3 | 9 | 274 | 9 | 1 | 731 | 130 |
| 2020-04-30 | 52 | 85 | 45 | 79 | 1091 | 7 | 6 | 275 | 17 | 1 | 748 | 131 |
| 2020-05-01 | 40 | 60 | 36 | 51 | 1076 | 4 | 9 | 270 | 11 | 0 | 759 | 131 |
| 2020-05-02 | 40 | 52 | 36 | 45 | 1067 | 4 | 7 | 267 | 10 | 5 | 769 | 136 |
| 2020-05-03 | 34 | 40 | 33 | 40 | 1060 | 1 | 0 | 268 | 17 | 0 | 786 | 136 |
| 2020-05-04 | 49 | 104 | 41 | 91 | 1010 | 8 | 13 | 263 | 9 | 2 | 795 | 138 |
| 2020-05-05 | 33 | 106 | 27 | 91 | 946 | 6 | 15 | 254 | 10 | 1 | 805 | 139 |
| 2020-05-06 | 41 | 86 | 39 | 69 | 916 | 2 | 17 | 239 | 10 | 0 | 815 | 139 |
| 2020-05-07 | 52 | 66 | 51 | 61 | 906 | 1 | 5 | 235 | 10 | 0 | 825 | 139 |
| 2020-05-08 | 29 | 88 | 25 | 68 | 863 | 4 | 20 | 219 | 7 | 3 | 832 | 142 |
| 2020-05-09 | 39 | 40 | 33 | 34 | 862 | 6 | 6 | 219 | 5 | 2 | 837 | 144 |
| 2020-05-10 | 34 | 26 | 30 | 22 | 870 | 4 | 4 | 219 | 9 | 3 | 846 | 147 |
| 2020-05-11 | 34 | 83 | 25 | 75 | 820 | 9 | 8 | 220 | 9 | 1 | 855 | 148 |
| 2020-05-12 | 38 | 67 | 35 | 56 | 799 | 3 | 11 | 212 | 7 | 2 | 862 | 150 |
| 2020-05-13 | 50 | 81 | 41 | 70 | 770 | 9 | 11 | 210 | 2 | 2 | 864 | 152 |
| 2020-05-14 | 49 | 73 | 46 | 60 | 756 | 3 | 13 | 200 | 4 | 2 | 868 | 154 |
| 2020-05-15 | 48 | 56 | 44 | 50 | 750 | 4 | 6 | 198 | 3 | 0 | 871 | 154 |
| 2020-05-16 | 37 | 38 | 31 | 34 | 747 | 6 | 4 | 200 | 9 | 2 | 880 | 156 |
| 2020-05-17 | 33 | 22 | 30 | 21 | 756 | 3 | 1 | 202 | 8 | 1 | 888 | 157 |
| 2020-05-18 | 25 | 74 | 24 | 66 | 714 | 1 | 8 | 195 | 3 | 3 | 891 | 160 |
| 2020-05-19 | 31 | 65 | 27 | 55 | 686 | 4 | 10 | 189 | 9 | 1 | 900 | 161 |
| 2020-05-20 | 44 | 72 | 39 | 62 | 663 | 5 | 10 | 184 | 7 | 0 | 907 | 161 |
| 2020-05-21 | 21 | 42 | 20 | 35 | 648 | 1 | 7 | 178 | 3 | 1 | 910 | 162 |
| 2020-05-22 | 36 | 83 | 34 | 70 | 612 | 2 | 13 | 167 | 1 | 3 | 911 | 165 |
| 2020-05-23 | 37 | 35 | 31 | 30 | 613 | 6 | 5 | 168 | 7 | 2 | 918 | 167 |
| 2020-05-24 | 34 | 15 | 28 | 12 | 629 | 6 | 3 | 171 | 3 | 2 | 921 | 169 |
| 2020-05-25 | 33 | 57 | 30 | 46 | 613 | 3 | 11 | 163 | 2 | 1 | 923 | 170 |
| 2020-05-26 | 39 | 46 | 37 | 37 | 613 | 2 | 9 | 156 | 5 | 2 | 928 | 172 |
| 2020-05-27 | 66 | 66 | 61 | 62 | 612 | 5 | 4 | 157 | 7 | 0 | 935 | 172 |
| 2020-05-28 | 55 | 60 | 49 | 55 | 606 | 6 | 5 | 158 | 6 | 2 | 941 | 174 |
| 2020-05-29 | 38 | 73 | 36 | 65 | 577 | 2 | 8 | 152 | 9 | 1 | 950 | 175 |
| 2020-05-30 | 33 | 25 | 30 | 25 | 582 | 3 | 0 | 155 | 5 | 0 | 955 | 175 |
| 2020-05-31 | 16 | 18 | 14 | 18 | 578 | 2 | 0 | 157 | 5 | 0 | 960 | 175 |

**Table 5:** Table with daily admissions (Part II)

| Date | Entries | Discharges | non-ICU<br>entries | non-ICU<br>dis-<br>charges | Non-ICU<br>in hospi-<br>tal | ICU en-<br>tries | ICU exits | ICU in<br>hospital | Non-ICU<br>deaths | ICU<br>deaths | sum<br>non-ICU<br>deaths | sum ICU<br>deaths |
| --- | --- | --- | --- | --- | --- | --- | --- | --- | --- | --- | --- | --- |
| 2020-06-01 | 49 | 54 | 41 | 47 | 572 | 8 | 7 | 158 | 9 | 2 | 969 | 177 |
| 2020-06-02 | 40 | 45 | 36 | 40 | 568 | 4 | 5 | 157 | 1 | 0 | 970 | 177 |
| 2020-06-03 | 42 | 51 | 38 | 44 | 562 | 4 | 7 | 154 | 4 | 2 | 974 | 179 |
| 2020-06-04 | 40 | 56 | 38 | 49 | 551 | 2 | 7 | 149 | 5 | 0 | 979 | 179 |
| 2020-06-05 | 35 | 65 | 31 | 56 | 526 | 4 | 9 | 144 | 11 | 1 | 990 | 180 |
| 2020-06-06 | 27 | 32 | 26 | 30 | 522 | 1 | 2 | 143 | 3 | 1 | 993 | 181 |
| 2020-06-07 | 28 | 18 | 21 | 17 | 526 | 7 | 1 | 149 | 4 | 0 | 997 | 181 |
| 2020-06-08 | 35 | 43 | 29 | 39 | 516 | 6 | 4 | 151 | 4 | 0 | 1001 | 181 |
| 2020-06-09 | 37 | 45 | 33 | 43 | 506 | 4 | 2 | 153 | 4 | 0 | 1005 | 181 |
| 2020-06-10 | 26 | 31 | 25 | 26 | 505 | 1 | 5 | 149 | 7 | 1 | 1012 | 182 |
| 2020-06-11 | 35 | 7 | 29 | 7 | 527 | 6 | 0 | 155 | 0 | 0 | 1012 | 182 |
| 2020-06-12 | 40 | 64 | 36 | 58 | 505 | 4 | 6 | 153 | 3 | 5 | 1015 | 187 |
| 2020-06-13 | 39 | 22 | 35 | 20 | 520 | 4 | 2 | 155 | 2 | 2 | 1017 | 189 |
| 2020-06-14 | 23 | 16 | 21 | 15 | 526 | 2 | 1 | 156 | 0 | 0 | 1017 | 189 |
| 2020-06-15 | 51 | 63 | 43 | 50 | 519 | 8 | 13 | 151 | 6 | 1 | 1023 | 190 |
| 2020-06-16 | 29 | 41 | 26 | 38 | 507 | 3 | 3 | 151 | 2 | 1 | 1025 | 191 |
| 2020-06-17 | 36 | 57 | 31 | 45 | 493 | 5 | 12 | 144 | 5 | 2 | 1030 | 193 |
| 2020-06-18 | 46 | 46 | 42 | 40 | 495 | 4 | 6 | 142 | 7 | 1 | 1037 | 194 |
| 2020-06-19 | 46 | 62 | 42 | 51 | 486 | 4 | 11 | 135 | 1 | 0 | 1038 | 194 |
| 2020-06-20 | 47 | 23 | 41 | 19 | 508 | 6 | 4 | 137 | 5 | 1 | 1043 | 195 |
| 2020-06-21 | 39 | 9 | 34 | 8 | 534 | 5 | 1 | 141 | 3 | 1 | 1046 | 196 |
| 2020-06-22 | 53 | 64 | 48 | 53 | 529 | 5 | 11 | 135 | 3 | 3 | 1049 | 199 |
| 2020-06-23 | 53 | 53 | 47 | 46 | 530 | 6 | 7 | 134 | 3 | 1 | 1052 | 200 |
| 2020-06-24 | 53 | 42 | 45 | 39 | 536 | 8 | 3 | 139 | 5 | 2 | 1057 | 202 |
| 2020-06-25 | 46 | 51 | 39 | 47 | 528 | 7 | 4 | 142 | 5 | 1 | 1062 | 203 |
| 2020-06-26 | 45 | 52 | 38 | 45 | 521 | 7 | 7 | 142 | 3 | 3 | 1065 | 206 |
| 2020-06-27 | 38 | 25 | 35 | 23 | 533 | 3 | 2 | 143 | 6 | 0 | 1071 | 206 |
| 2020-06-28 | 37 | 13 | 31 | 12 | 552 | 6 | 1 | 148 | 4 | 0 | 1075 | 206 |
| 2020-06-29 | 37 | 62 | 29 | 54 | 527 | 8 | 8 | 148 | 7 | 2 | 1082 | 208 |
| 2020-06-30 | 31 | 36 | 27 | 32 | 522 | 4 | 4 | 148 | 5 | 0 | 1087 | 208 |
| 2020-07-01 | 39 | 59 | 34 | 50 | 506 | 5 | 9 | 144 | 5 | 3 | 1092 | 211 |
| 2020-07-02 | 33 | 46 | 31 | 42 | 495 | 2 | 4 | 142 | 7 | 4 | 1099 | 215 |
| 2020-07-03 | 51 | 65 | 45 | 61 | 479 | 6 | 4 | 144 | 8 | 1 | 1107 | 216 |
| 2020-07-04 | 37 | 23 | 33 | 18 | 494 | 4 | 5 | 143 | 7 | 1 | 1114 | 217 |
| 2020-07-05 | 37 | 22 | 31 | 19 | 506 | 6 | 3 | 146 | 5 | 2 | 1119 | 219 |
| 2020-07-06 | 44 | 59 | 39 | 54 | 491 | 5 | 5 | 146 | 7 | 1 | 1126 | 220 |
| 2020-07-07 | 39 | 37 | 35 | 31 | 495 | 4 | 6 | 144 | 5 | 3 | 1131 | 223 |
| 2020-07-08 | 40 | 58 | 37 | 48 | 484 | 3 | 10 | 137 | 11 | 2 | 1142 | 225 |
| 2020-07-09 | 35 | 52 | 32 | 46 | 470 | 3 | 6 | 134 | 3 | 0 | 1145 | 225 |
| 2020-07-10 | 40 | 45 | 34 | 38 | 466 | 6 | 7 | 133 | 7 | 2 | 1152 | 227 |
| 2020-07-11 | 23 | 25 | 23 | 18 | 471 | 0 | 7 | 126 | 5 | 5 | 1157 | 232 |
| 2020-07-12 | 25 | 8 | 21 | 8 | 484 | 4 | 0 | 130 | 1 | 0 | 1158 | 232 |
| 2020-07-13 | 36 | 38 | 32 | 29 | 487 | 4 | 9 | 125 | 4 | 2 | 1162 | 234 |
| 2020-07-14 | 30 | 42 | 29 | 39 | 477 | 1 | 3 | 123 | 6 | 0 | 1168 | 234 |
| 2020-07-15 | 36 | 39 | 30 | 34 | 473 | 6 | 5 | 124 | 4 | 0 | 1172 | 234 |
| 2020-07-16 | 29 | 37 | 25 | 31 | 467 | 4 | 6 | 122 | 3 | 0 | 1175 | 234 |
| 2020-07-17 | 30 | 51 | 24 | 42 | 449 | 6 | 9 | 119 | 3 | 0 | 1178 | 234 |
| 2020-07-18 | 26 | 20 | 24 | 17 | 456 | 2 | 3 | 118 | 5 | 1 | 1183 | 235 |
| 2020-07-19 | 28 | 13 | 24 | 12 | 468 | 4 | 1 | 121 | 4 | 1 | 1187 | 236 |
| 2020-07-20 | 32 | 44 | 27 | 39 | 456 | 5 | 5 | 121 | 4 | 3 | 1191 | 239 |
| 2020-07-21 | 29 | 39 | 27 | 31 | 452 | 2 | 8 | 115 | 5 | 2 | 1196 | 241 |
| 2020-07-22 | 27 | 33 | 25 | 30 | 447 | 2 | 3 | 114 | 3 | 1 | 1199 | 242 |
| 2020-07-23 | 33 | 52 | 29 | 43 | 433 | 4 | 9 | 109 | 5 | 2 | 1204 | 244 |
| 2020-07-24 | 32 | 47 | 28 | 39 | 422 | 4 | 8 | 105 | 3 | 0 | 1207 | 244 |
| 2020-07-25 | 24 | 18 | 20 | 16 | 426 | 4 | 2 | 107 | 3 | 0 | 1210 | 244 |
| 2020-07-26 | 28 | 12 | 28 | 10 | 444 | 0 | 2 | 105 | 2 | 1 | 1212 | 245 |
| 2020-07-27 | 19 | 34 | 18 | 32 | 430 | 1 | 2 | 104 | 2 | 0 | 1214 | 245 |
| 2020-07-28 | 23 | 30 | 19 | 26 | 423 | 4 | 4 | 104 | 2 | 1 | 1216 | 246 |
| 2020-07-29 | 22 | 35 | 19 | 30 | 412 | 3 | 5 | 102 | 4 | 0 | 1220 | 246 |
| 2020-07-30 | 21 | 42 | 18 | 36 | 394 | 3 | 6 | 99 | 5 | 3 | 1225 | 249 |
| 2020-07-31 | 20 | 58 | 18 | 45 | 367 | 2 | 13 | 88 | 1 | 1 | 1226 | 250 |
| 2020-08-01 | 22 | 9 | 18 | 9 | 376 | 4 | 0 | 92 | 1 | 0 | 1227 | 250 |
| 2020-08-02 | 22 | 6 | 21 | 4 | 393 | 1 | 2 | 91 | 1 | 2 | 1228 | 252 |
| 2020-08-03 | 15 | 28 | 12 | 24 | 381 | 3 | 4 | 90 | 1 | 0 | 1229 | 252 |
| 2020-08-04 | 16 | 29 | 13 | 26 | 368 | 3 | 3 | 90 | 3 | 0 | 1232 | 252 |
| 2020-08-05 | 20 | 27 | 18 | 23 | 363 | 2 | 4 | 88 | 4 | 0 | 1236 | 252 |
| 2020-08-06 | 21 | 32 | 19 | 27 | 355 | 2 | 5 | 85 | 6 | 1 | 1242 | 253 |
| 2020-08-07 | 43 | 49 | 42 | 41 | 356 | 1 | 8 | 78 | 1 | 3 | 1243 | 256 |
| 2020-08-08 | 23 | 16 | 21 | 15 | 362 | 2 | 1 | 79 | 4 | 1 | 1247 | 257 |
| 2020-08-09 | 24 | 12 | 21 | 10 | 373 | 3 | 2 | 80 | 4 | 1 | 1251 | 258 |
| 2020-08-10 | 29 | 37 | 24 | 36 | 361 | 5 | 1 | 84 | 2 | 0 | 1253 | 258 |
| 2020-08-11 | 29 | 19 | 25 | 17 | 369 | 4 | 2 | 86 | 3 | 0 | 1256 | 258 |
| 2020-08-12 | 16 | 25 | 15 | 20 | 364 | 1 | 5 | 82 | 4 | 0 | 1260 | 258 |
| 2020-08-13 | 13 | 27 | 9 | 22 | 351 | 4 | 5 | 81 | 5 | 0 | 1265 | 258 |
| 2020-08-14 | 18 | 27 | 15 | 24 | 342 | 3 | 3 | 81 | 3 | 0 | 1268 | 258 |
| 2020-08-15 | 25 | 13 | 18 | 12 | 348 | 7 | 1 | 87 | 4 | 1 | 1272 | 259 |
| 2020-08-16 | 20 | 6 | 18 | 6 | 360 | 2 | 0 | 89 | 2 | 0 | 1274 | 259 |
| 2020-08-17 | 27 | 33 | 22 | 25 | 357 | 5 | 8 | 86 | 4 | 2 | 1278 | 261 |
| 2020-08-18 | 20 | 28 | 20 | 26 | 351 | 0 | 2 | 84 | 1 | 1 | 1279 | 262 |
| 2020-08-19 | 29 | 29 | 21 | 25 | 347 | 8 | 4 | 88 | 4 | 0 | 1283 | 262 |
| 2020-08-20 | 25 | 32 | 22 | 30 | 339 | 3 | 2 | 89 | 2 | 1 | 1285 | 263 |
| 2020-08-21 | 22 | 14 | 19 | 12 | 346 | 3 | 2 | 90 | 2 | 0 | 1287 | 263 |
| 2020-08-22 | 20 | 12 | 18 | 11 | 353 | 2 | 1 | 91 | 4 | 0 | 1291 | 263 |
| 2020-08-23 | 25 | 17 | 18 | 14 | 357 | 7 | 3 | 95 | 4 | 2 | 1295 | 265 |
| 2020-08-24 | 23 | 24 | 20 | 19 | 358 | 3 | 5 | 93 | 3 | 2 | 1298 | 267 |
| 2020-08-25 | 25 | 34 | 21 | 30 | 349 | 4 | 4 | 93 | 8 | 0 | 1306 | 267 |
| 2020-08-26 | 31 | 37 | 27 | 35 | 341 | 4 | 2 | 95 | 2 | 0 | 1308 | 267 |
| 2020-08-27 | 36 | 25 | 28 | 19 | 350 | 8 | 6 | 97 | 4 | 1 | 1312 | 268 |
| 2020-08-28 | 32 | 42 | 27 | 35 | 342 | 5 | 7 | 95 | 1 | 0 | 1313 | 268 |
| 2020-08-29 | 30 | 12 | 26 | 12 | 356 | 4 | 0 | 99 | 3 | 0 | 1316 | 268 |
| 2020-08-30 | 23 | 12 | 20 | 12 | 364 | 3 | 0 | 102 | 4 | 0 | 1320 | 268 |
| 2020-08-31 | 29 | 30 | 24 | 25 | 363 | 5 | 5 | 102 | 2 | 0 | 1322 | 268 |

**Table 6:**
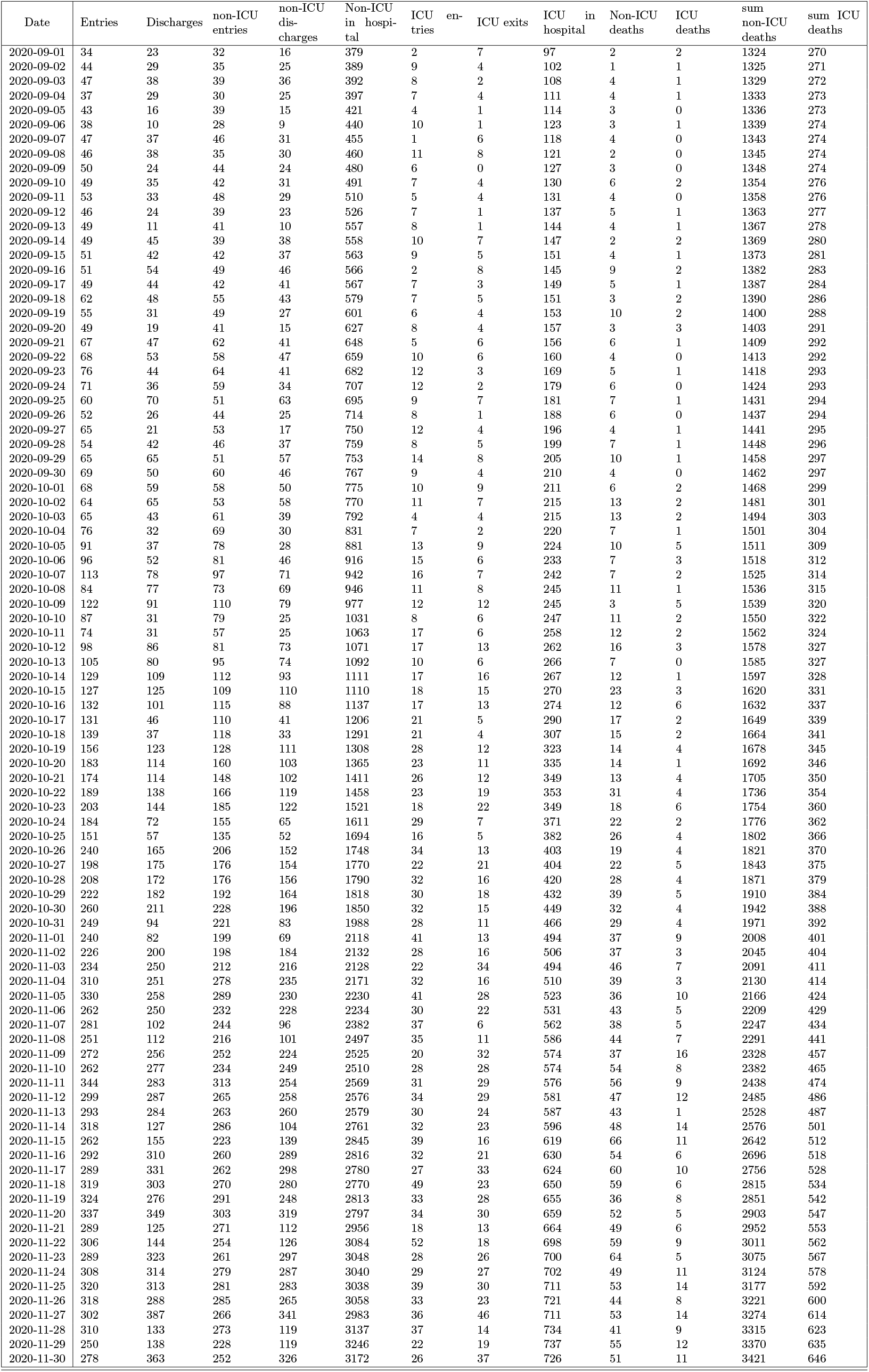
Table with daily admissions (Pare III)

**Table 7:**
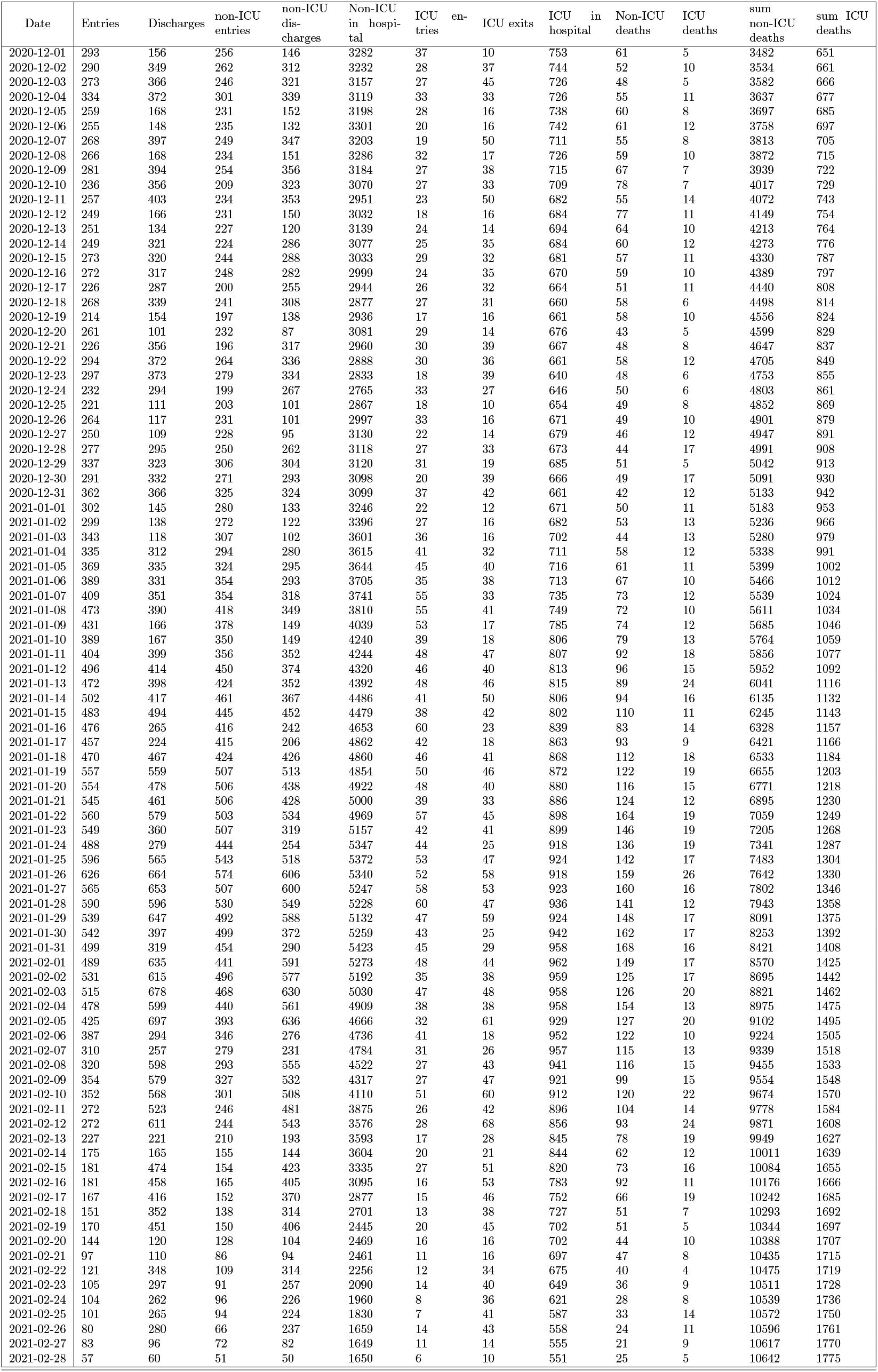
Table with dmissions aggregataed by day (Part IV)

**Table 8:** Table with daily admissions (Part 5)

| Date | Entries | Discharges | non-ICU entries | non-ICU discharges | Non-ICU in hospital | ICU entries | ICU exits | ICU in hospital | Non-ICU deaths | ICU deaths | sum non-ICU deaths | sum ICU deaths |
| --- | --- | --- | --- | --- | --- | --- | --- | --- | --- | --- | --- | --- |
| 2021-03-01 | 86 | 244 | 80 | 203 | 1527 | 6 | 41 | 516 | 23 | 11 | 10665 | 1786 |
| 2021-03-02 | 70 | 200 | 58 | 172 | 1413 | 12 | 28 | 500 | 30 | 5 | 10695 | 1791 |
| 2021-03-03 | 71 | 181 | 63 | 156 | 1320 | 8 | 25 | 483 | 22 | 4 | 10717 | 1795 |
| 2021-03-04 | 67 | 176 | 57 | 153 | 1224 | 10 | 23 | 470 | 24 | 5 | 10741 | 1800 |
| 2021-03-05 | 68 | 209 | 64 | 180 | 1108 | 4 | 29 | 445 | 18 | 5 | 10759 | 1805 |
| 2021-03-06 | 60 | 53 | 55 | 45 | 1118 | 5 | 8 | 442 | 19 | 6 | 10778 | 1811 |
| 2021-03-07 | 38 | 41 | 35 | 32 | 1121 | 3 | 9 | 436 | 17 | 5 | 10795 | 1816 |
| 2021-03-08 | 45 | 184 | 40 | 155 | 1006 | 5 | 29 | 412 | 17 | 11 | 10812 | 1827 |
| 2021-03-09 | 52 | 149 | 43 | 123 | 926 | 9 | 26 | 395 | 13 | 6 | 10825 | 1833 |
| 2021-03-10 | 50 | 127 | 47 | 109 | 864 | 3 | 18 | 380 | 12 | 3 | 10837 | 1836 |
| 2021-03-11 | 41 | 112 | 35 | 91 | 808 | 6 | 21 | 365 | 10 | 4 | 10847 | 1840 |
| 2021-03-12 | 33 | 132 | 28 | 111 | 725 | 5 | 21 | 349 | 16 | 3 | 10863 | 1843 |
| 2021-03-13 | 33 | 39 | 30 | 32 | 723 | 3 | 7 | 345 | 7 | 7 | 10870 | 1850 |
| 2021-03-14 | 27 | 27 | 25 | 22 | 726 | 2 | 5 | 342 | 11 | 4 | 10881 | 1854 |
| 2021-03-15 | 29 | 95 | 28 | 71 | 683 | 1 | 24 | 319 | 6 | 6 | 10887 | 1860 |
| 2021-03-16 | 38 | 111 | 33 | 89 | 627 | 5 | 22 | 302 | 9 | 5 | 10896 | 1865 |
| 2021-03-17 | 41 | 82 | 35 | 65 | 597 | 6 | 17 | 291 | 6 | 7 | 10902 | 1872 |
| 2021-03-18 | 39 | 79 | 37 | 59 | 575 | 2 | 20 | 273 | 6 | 3 | 10908 | 1875 |
| 2021-03-19 | 35 | 100 | 31 | 73 | 533 | 4 | 27 | 250 | 8 | 5 | 10916 | 1880 |
| 2021-03-20 | 24 | 18 | 22 | 11 | 544 | 2 | 7 | 245 | 3 | 3 | 10919 | 1883 |
| 2021-03-21 | 28 | 24 | 24 | 18 | 550 | 4 | 6 | 243 | 6 | 5 | 10925 | 1888 |
| 2021-03-22 | 40 | 76 | 36 | 65 | 521 | 4 | 11 | 236 | 5 | 1 | 10930 | 1889 |
| 2021-03-23 | 37 | 79 | 32 | 57 | 496 | 5 | 22 | 219 | 6 | 7 | 10936 | 1896 |
| 2021-03-24 | 34 | 63 | 31 | 56 | 471 | 3 | 7 | 215 | 5 | 2 | 10941 | 1898 |
| 2021-03-25 | 34 | 60 | 29 | 51 | 449 | 5 | 9 | 211 | 5 | 0 | 10946 | 1898 |
| 2021-03-26 | 32 | 80 | 28 | 62 | 415 | 4 | 18 | 197 | 3 | 3 | 10949 | 1901 |
| 2021-03-27 | 25 | 25 | 22 | 20 | 417 | 3 | 5 | 195 | 4 | 4 | 10953 | 1905 |
| 2021-03-28 | 13 | 21 | 10 | 18 | 409 | 3 | 3 | 195 | 5 | 2 | 10958 | 1907 |
| 2021-03-29 | 22 | 63 | 20 | 56 | 373 | 2 | 7 | 190 | 2 | 1 | 10960 | 1908 |
| 2021-03-30 | 25 | 50 | 23 | 35 | 361 | 2 | 15 | 177 | 4 | 1 | 10964 | 1909 |
| 2021-03-31 | 34 | 61 | 32 | 48 | 345 | 2 | 13 | 166 | 8 | 3 | 10972 | 1912 |

**Table 9:**
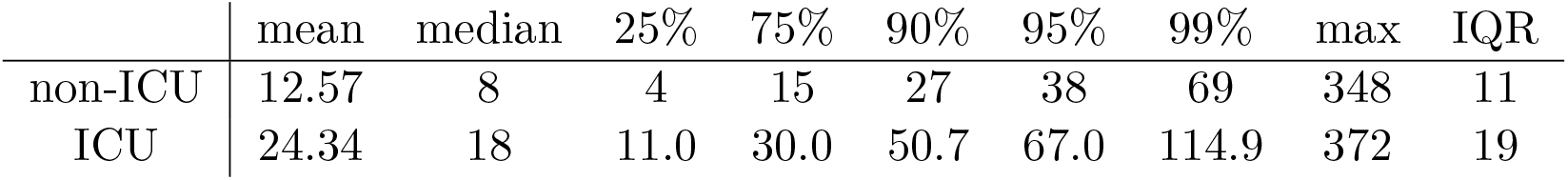
Summary statistics of the LoS before data trimming.

Summary statistics complementary to the various Violin/box plots and histograms are available in appendix A.4. Extra graphs of interest may be found in appendix A.5. All data processing and analysis was done using *R* [14].

### 2.2. Distribution fitting

The probability distributions of the LoS and TuD, for non-ICU and ICU patients, were also studied using the R *fitdistrplus* package [5]. The objective of this analysis was to find the probability distribution that best describes the observed data. To enable the use of the Weibull distributions the episodes with 0 days of stay were removed.

To choose the candidate distributions to be fitted to the data we used different tools of *fitdistrplus*. In addition to empirical plots, like density histograms and the plots of the empirical cumulative distribution function, the descriptive statistics provided helps in this choice, specially the Skewness-kurtosis plot (figure 11), also known as Cullen and Frey graph, with a nonparametric bootstrap procedure to take into account the uncertainty in the estimation of kurtosis and skewness from data.

The next step was to fit the chosen distributions to the data. The default method of estimation uses the maximum likelihood function, which was used in all chosen distributions to be fitted. With the objective to give more weight to the left tail of the distribution, as the data has many large outliers, the maximum goodness-of-fit estimation using the left-tail Anderson-Darling (ADL) distance was used in the two best fitted distributions, the gamma and log-normal distributions.

To evaluate the fit of the candidate distribution, various goodness-of-fit statistics (Kolmogorov-Smirnov, Cramer-von Mises and Anderson-Darling statistic) and goodness-of-fit criteria (Aikake’s and Bayesian information criterion) are provided in the appendix A.4.

## 3. RESULTS

### 3.1. Exploratory data analysis

One of the most important parameters related to hospitalisation is the length of stay (LoS), as the rate of exits from hospital and the number of occupied beds are dependent on it. In figure 1 a violin/box plot, together with histograms, related to LoS for either type of hospitalisation are presented, obtaining a mean of 12.5 days and a median of 8 days (IQR:4-15) for non-ICU patients and a mean of 24.3 days and median of 18 days (IQR:11-30) for ICU patients (extended summary statistics avaliable at appendix A.4 table 10). There is a clear difference in LoS between non-ICU and ICU patients.

**Figure 1:**
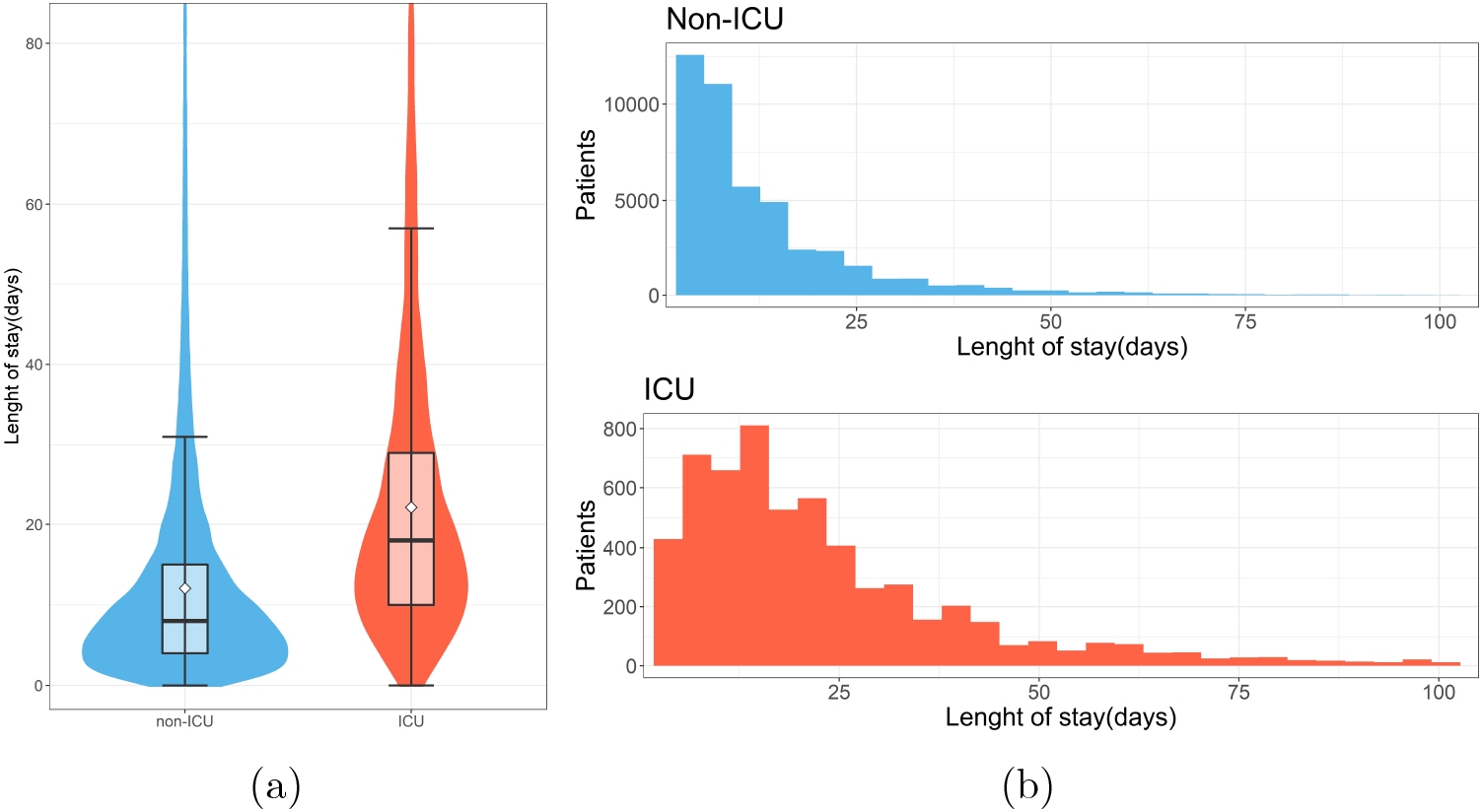
Distribution of the LoS in hospital care. **(a)** Violin/box plot of the length of stay by type of hospitalisation, in which the white diamond represents the mean. **(b)** Distribution of the LoS for non-ICU and ICU patients (white diamonds represent the mean).

**Table 10:** Summary statistics of the LoS by type of hospitalisation (non-ICU and ICU patients).

| LoS | mean | median | 25% | 75% | 90% | 95% | 99% | max | IQR |
| --- | --- | --- | --- | --- | --- | --- | --- | --- | --- |
| Non-ICU | 12.54 | 8 | 4 | 15 | 27 | 38 | 67 | 348 | 11 |
| ICU | 24.31 | 18 | 11.0 | 30.3 | 51.0 | 66.0 | 112.0 | 216 | 19.3 |

There are two possible hospitalisation outcomes, either the patient is discharged from hospital or he dies. Considering these outcomes a violin/box plot comparing time until death (TuD) and time until Discharge (TuDisc) is presented in figure 2, with summary statistics available in appendix A.4 in table 11. The TuD mean and median are 11.01 and 7 days for non-ICU, and 20.04 and 16 for ICU, respectively, and for TuDisc the mean and median are 12.98 and 9 for non-ICU and 26.39 and 19 days for ICU patient.

**Figure 2:**
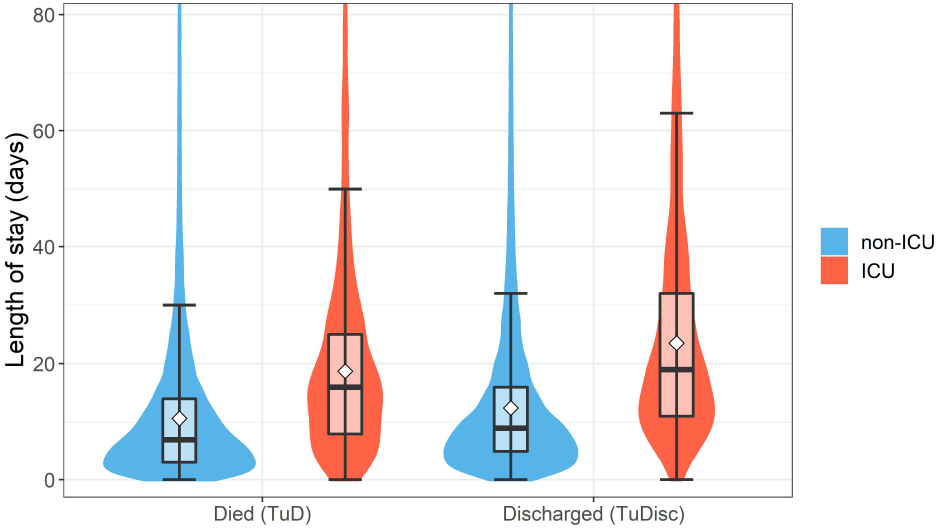
Violin/Box plot of the length of stay (LoS) for each outcome (Death or Discharge) and type of hospitalisation (non-ICU and ICU) (white diamonds represent the mean).

**Table 11:** Summary statistics of the length of stay for patients that died (TuD) or where discharged (TuDisc), for either type of hospitalisation (non-ICU and ICU patients).

|  |  | mean | median | 25% | 75% | 90% | 95% | 99% | max | IQR |
| --- | --- | --- | --- | --- | --- | --- | --- | --- | --- | --- |
| TuD | Non-ICU | 11.01 | 7 | 3 | 14 | 24 | 33 | 61 | 228 | 11 |
|  | ICU | 20.04 | 16 | 9.0 | 26.0 | 39.0 | 53.0 | 95.4 | 199.0 | 17 |
| TuDisc | Non-ICU | 12.98 | 9 | 5 | 16 | 28 | 39 | 69 | 348 | 11 |
|  | ICU | 26.39 | 19 | 12 | 34 | 56 | 71 | 115 | 216 | 22 |

Three parameters that describe the hospitalisation dynamics were estimated as follows:

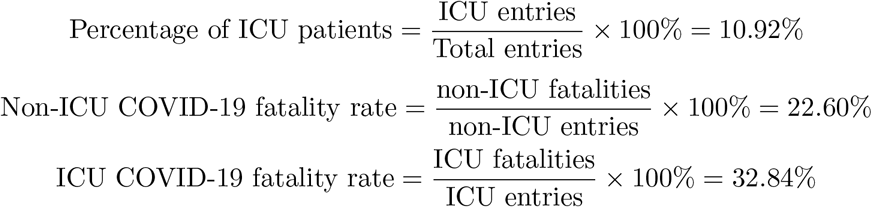

In which the percentage of ICU patients represents the percentage of patients that, at any point during hospital care, received ICU treatment, the non-ICU COVID-19 fatality rate represents the percentage of patients that did not receive any non-ICU care and that died in hospital care, and the ICU COVID-19 fatality rate represents the percentage of patients that at any point during hospital care received ICU treatment and died in hospital care.

#### 3.1.1. Characterization over time

One of the characteristics of the COVID-19 pandemic is the peak in cases that occurs in different periods of time. The number of hospitalizations follows the number of cases and therefore is also prone to rapid peaks in hospitalisation. This is specially problematic for health services since this could lead to a lack of hospital capacity and affect the care to be provided.

We found an association between number of hospitalizations and the fatality rate, the higher the number of hospital entries the higher the in-hospital fatality rate and the decrease in the LoS of the patients that died, as seen in figure 3. There is visible the increase in fatality rate with the increase in cases related to the second surge in hospitalisation from September to February 2020.

**Figure 3:**
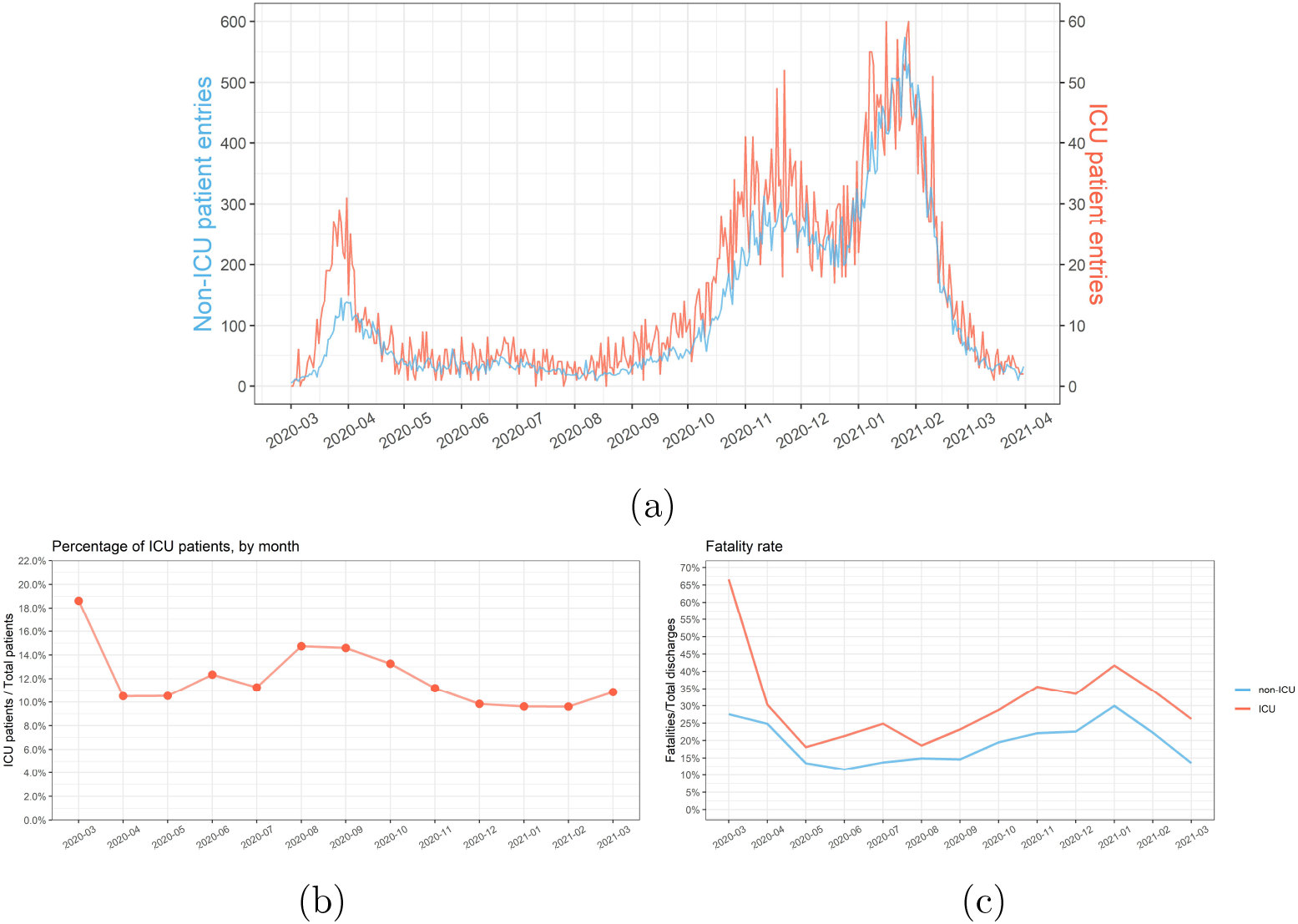
**(a)**Entries, **(b)**percentage of ICU patients and **(c)**fatality rate, over time and by type of hospitalisation

**Figure 4:**
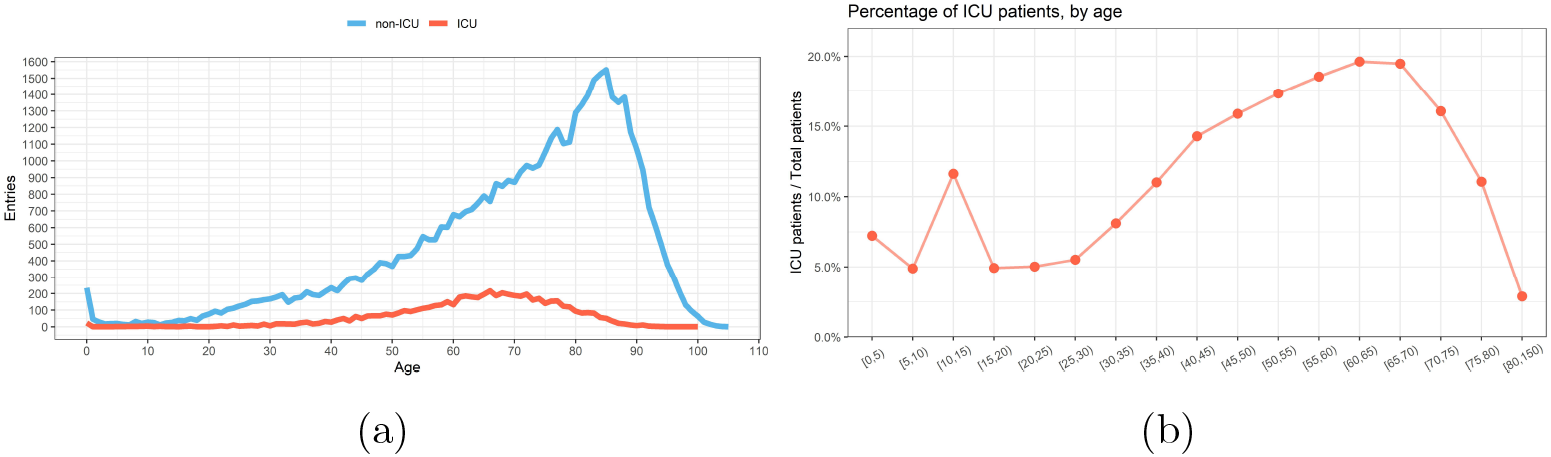
Hospital entries by age: **(a)**entries and **(b)**percentage of patients requiring ICU treatment.

**Figure 5:**
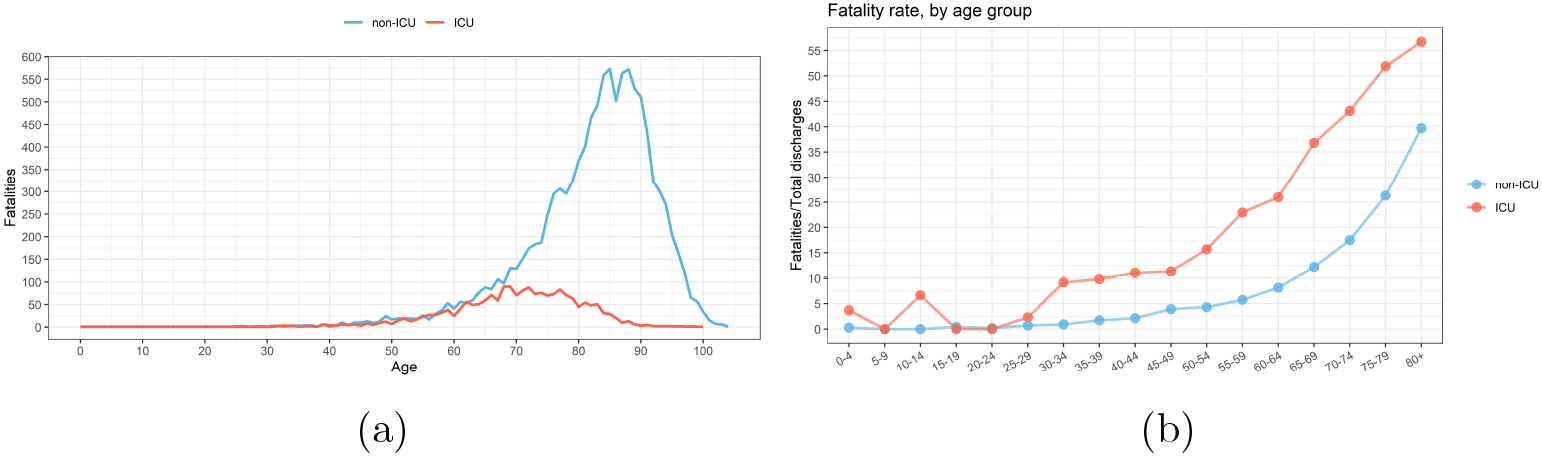
Mortality by age: **(a)**number of fatalities and **(b)**in-hospital fatality rate.

**Figure 6:**
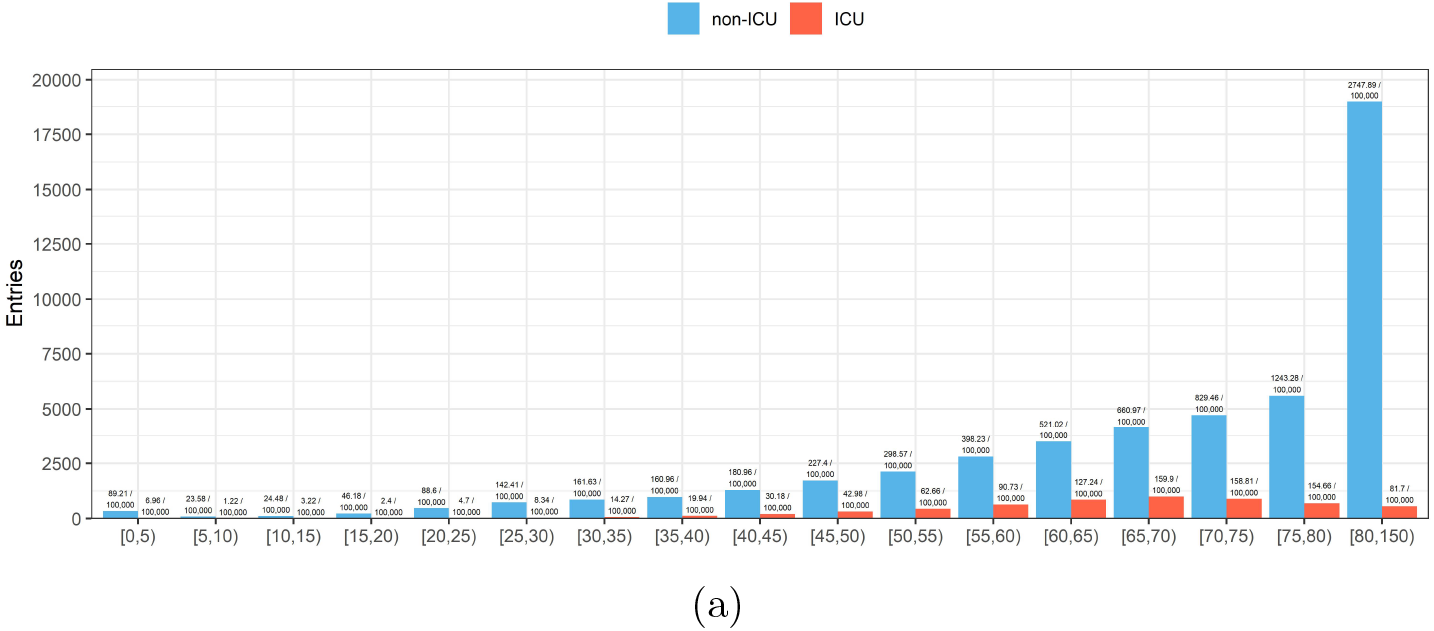
Hospital entries by age group and type of hospitalization.

**Figure 7:**
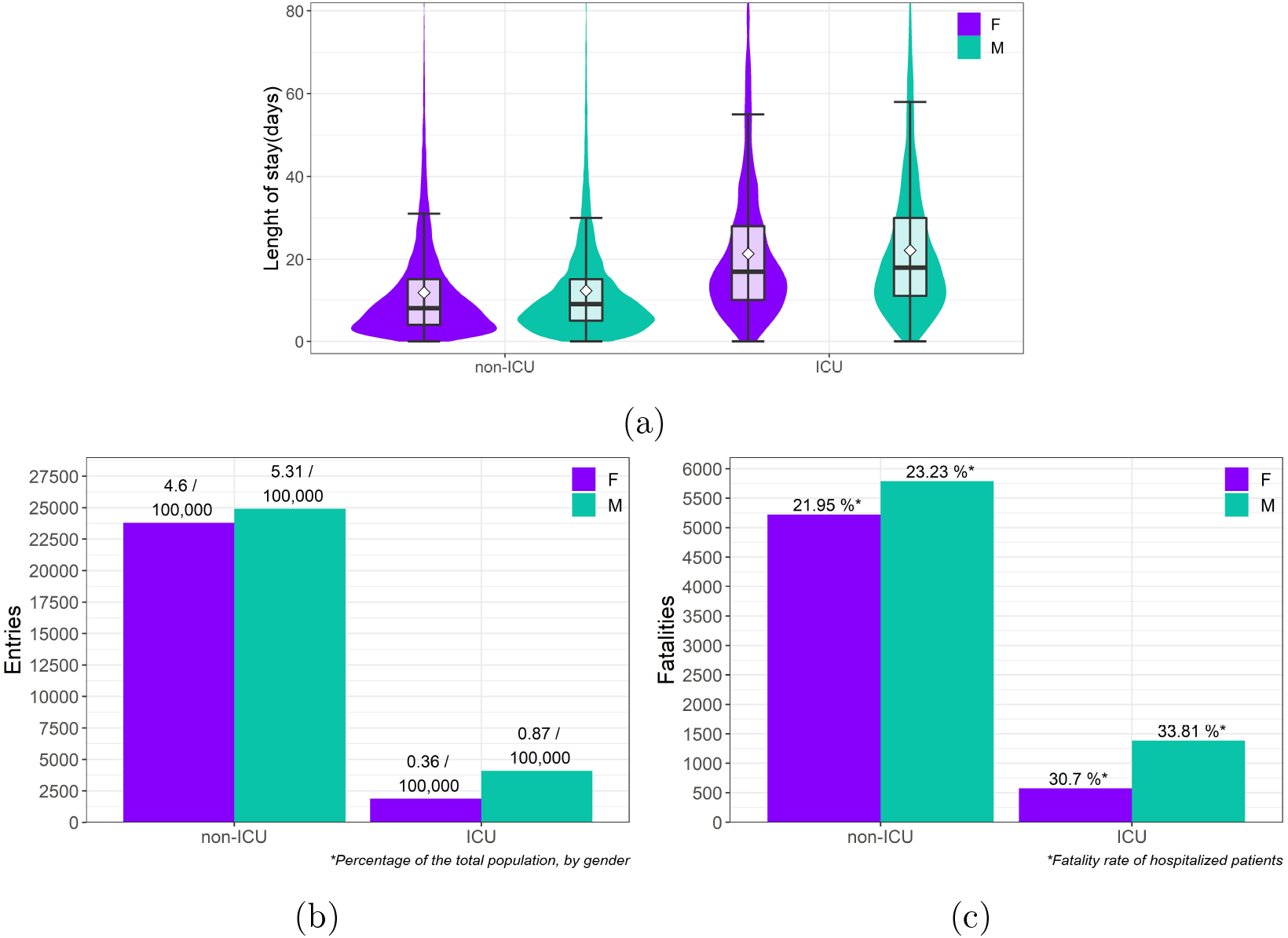
Hospital entries by gender (F for female and M for male, white diamonds represent the mean). (a)Violin/box plot of the LoS by gender and type of hospitalisation, (b)total number of entries by gender and type of hospitalisation and (c)total number of fatalities by gender and type of hospitalisation, with fatality rates.

**Figure 8:**
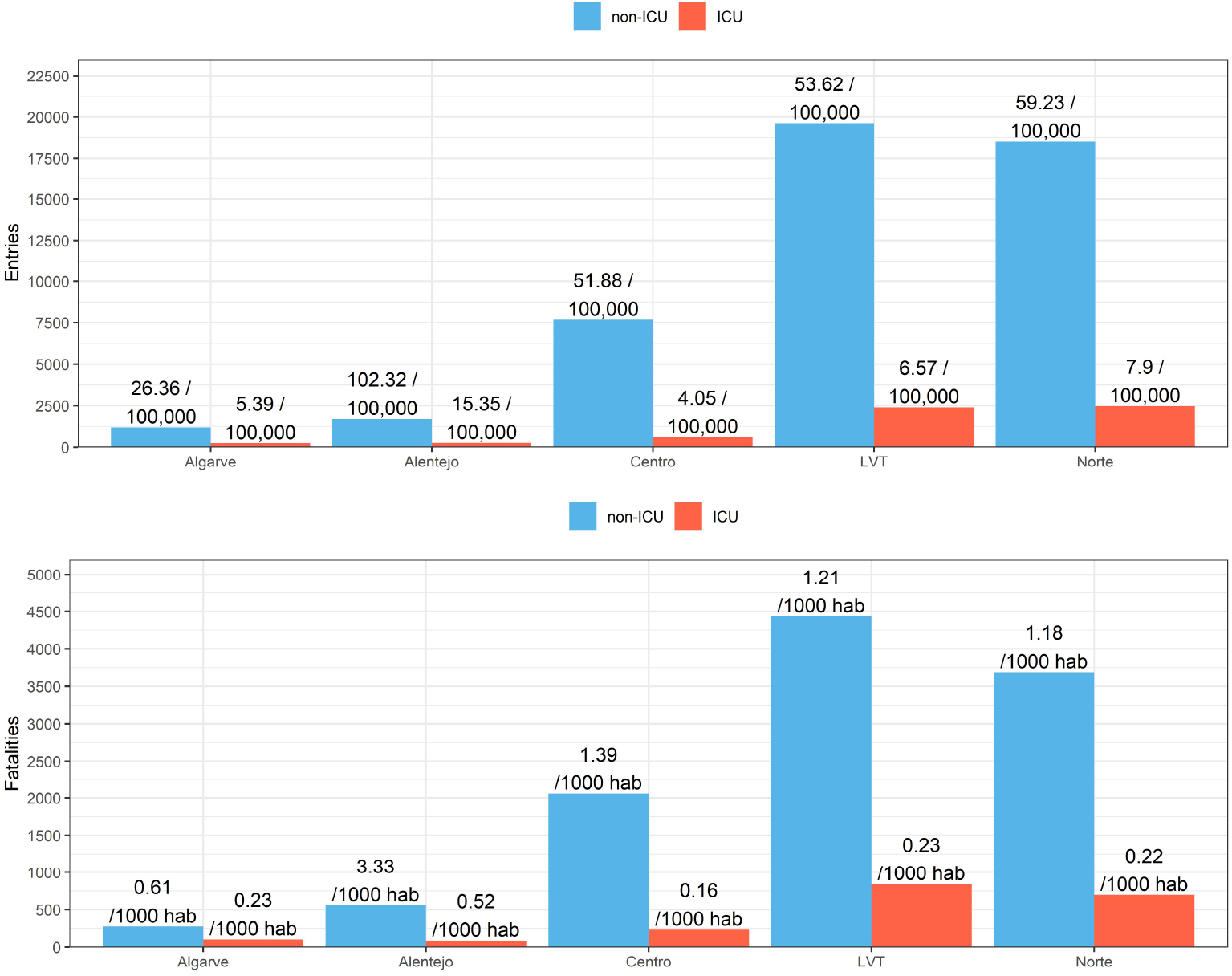
Number of (a)entries and (b)fatalities, by region and type of hospitalisation.

Violin/box plots for either LoS and TuD over time are available in appendix A.5, in figure 14 (a) and (b), linegraphs with hospital occupancy, discharges and fatalities over time are also available in figure 15. Summary statistics for LoS and TuD are displayed in appendix A.4 table 12 and 13.

**Figure 9:**
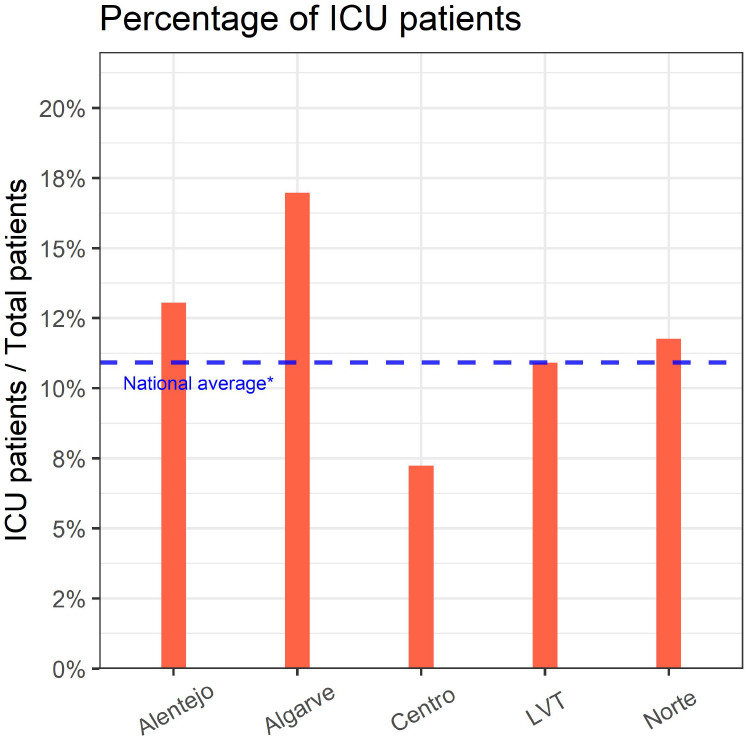

**Figure 10:** Percentage of ICU patients, by region.

**Figure 11:**
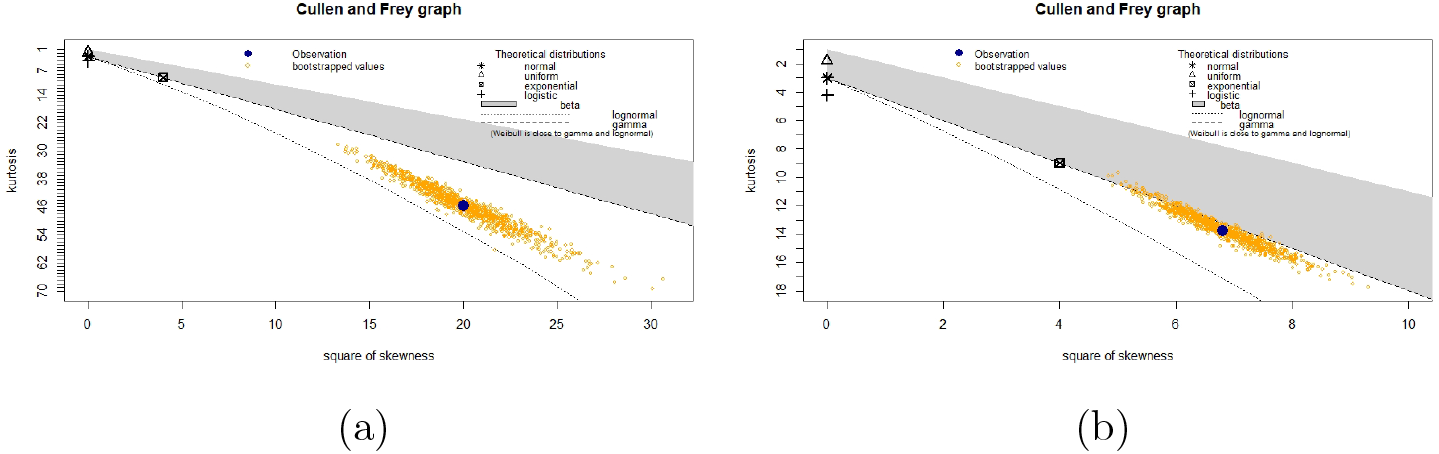
Cullen and Frey graphs for (a)Non-ICU and (b)ICU patients.

**Figure 12:**
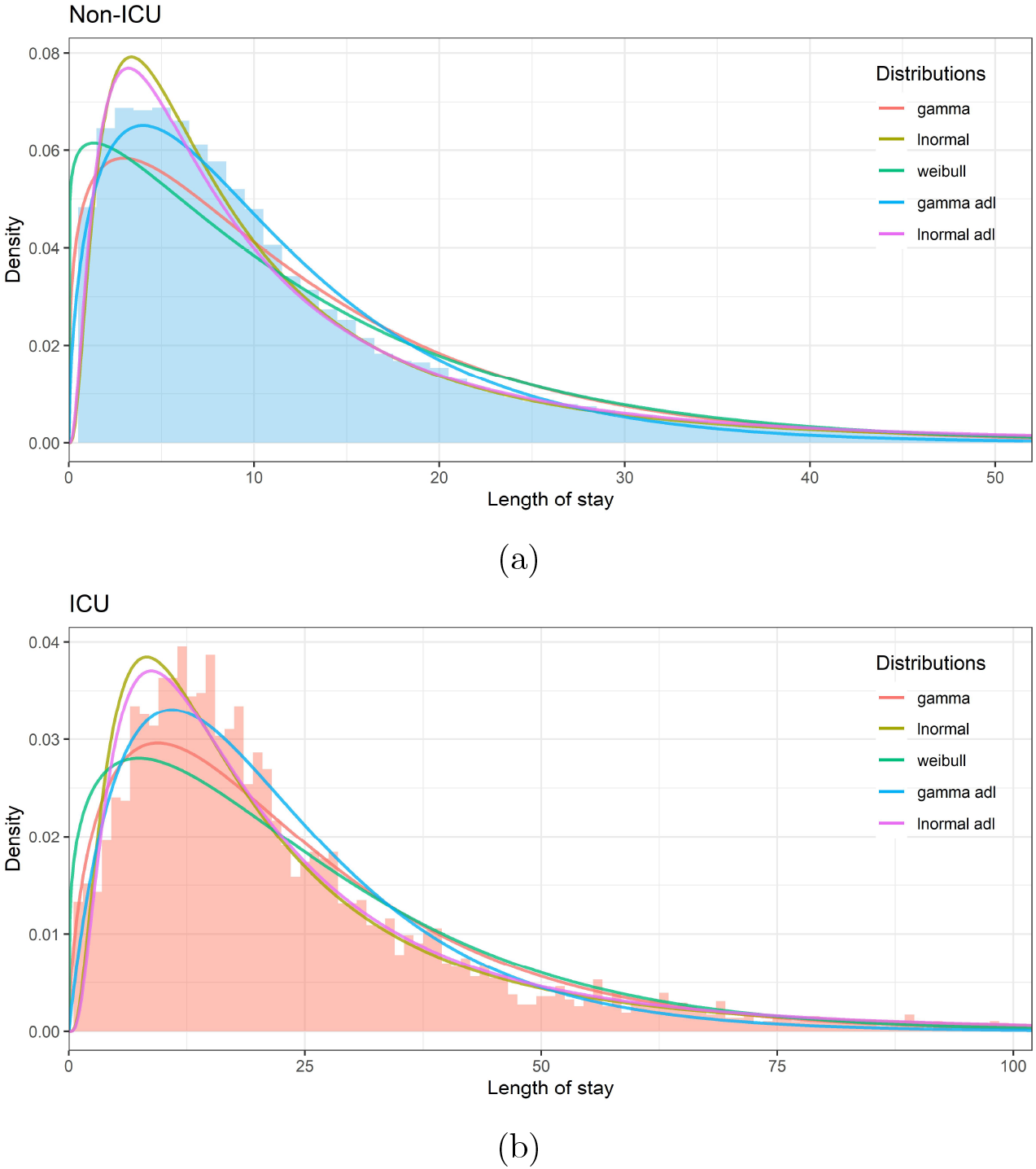
Fitted distributions to the LoS for (a)non-ICU (Hosp) and (b)ICU patients.

**Figure 13:**
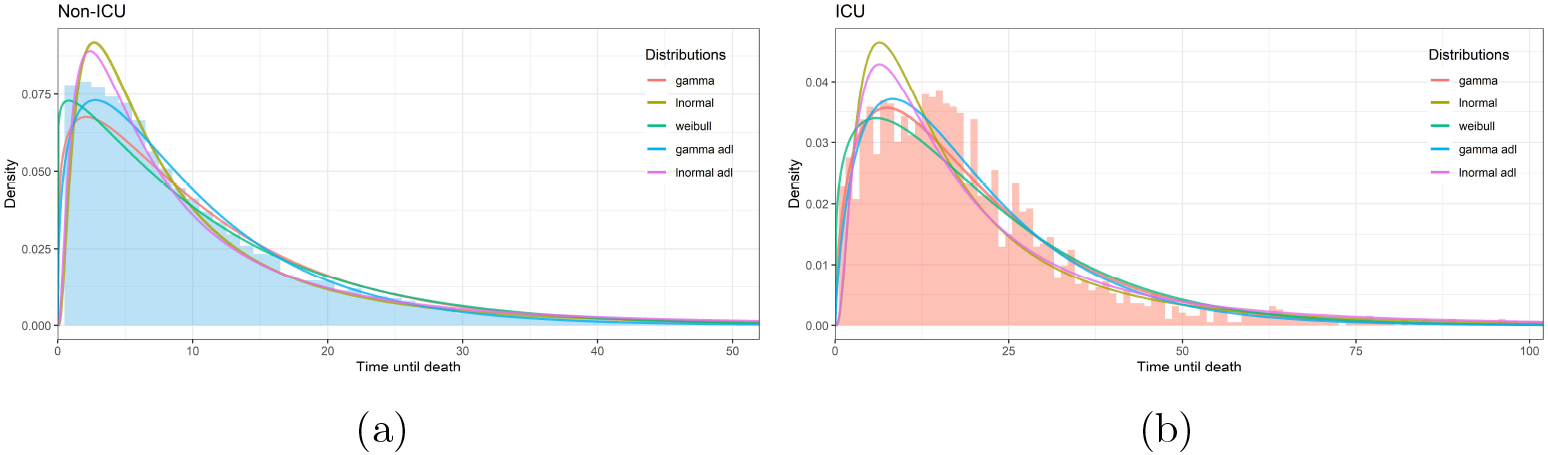
Fitted distributions to the time until death for (a)non-ICU and (b)ICU patients.

**Figure 14:**
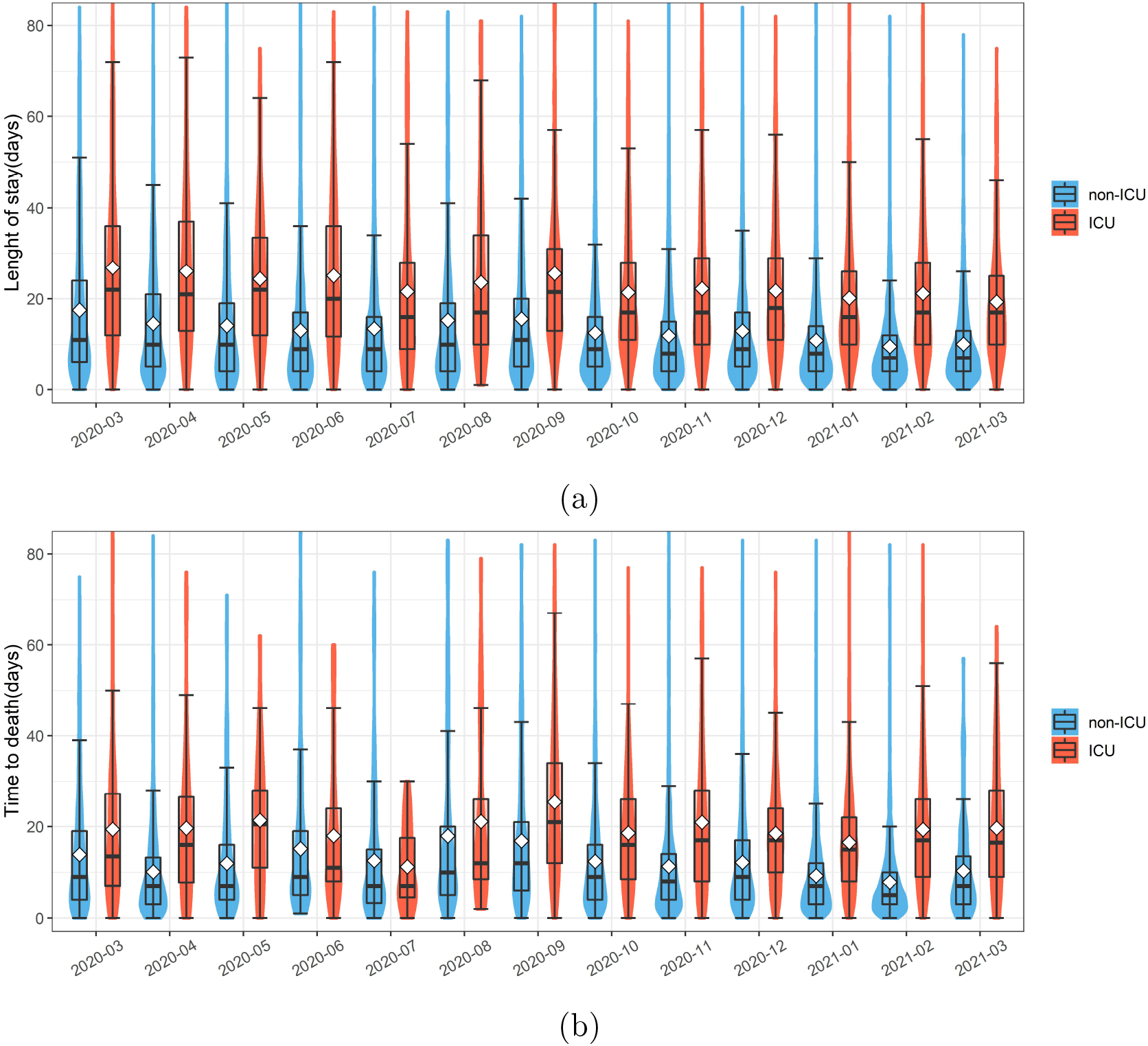
Violin/box plots of the **(a)**LoS and **(b)**TuD, by month and type of hospitalisation (white diamonds represent the mean).

**Figure 15:**
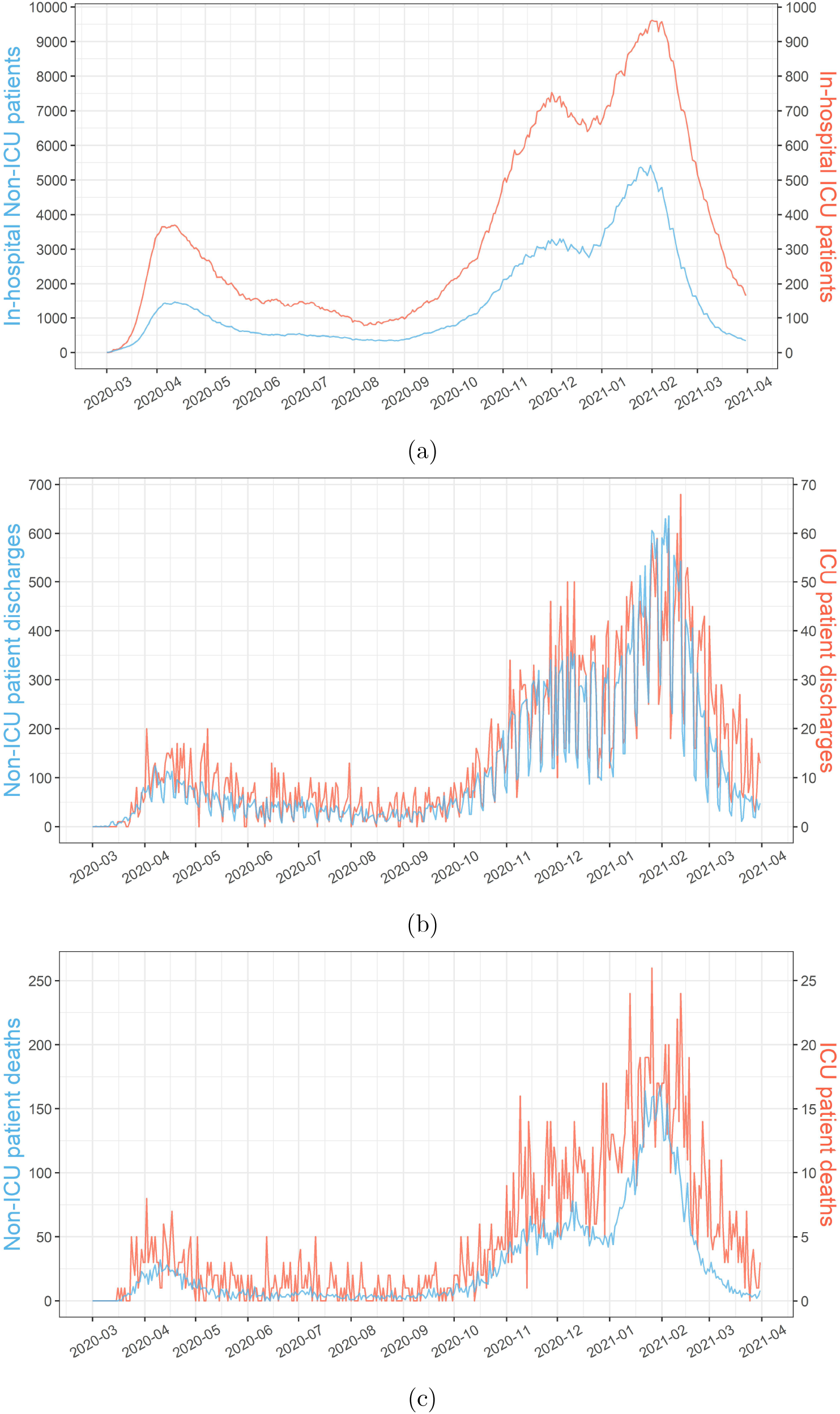
Evolution of patients (a)in-hospital, (b)discharged and (c)death with time, by type of hospitalization.

**Table 12:**
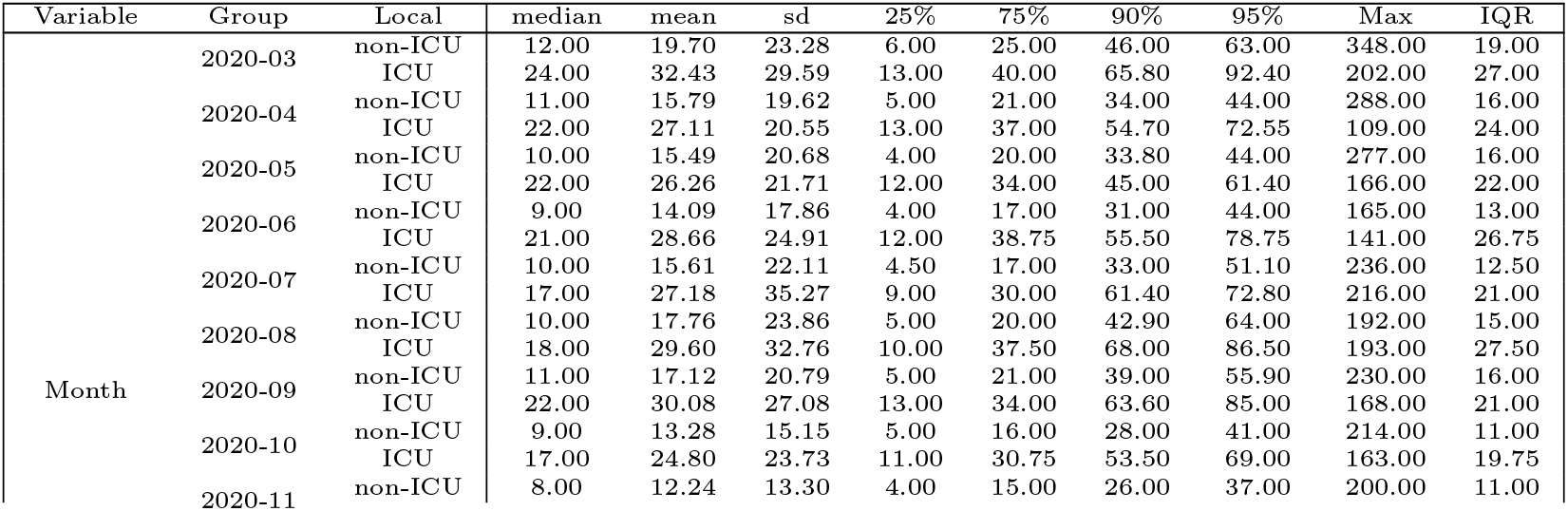
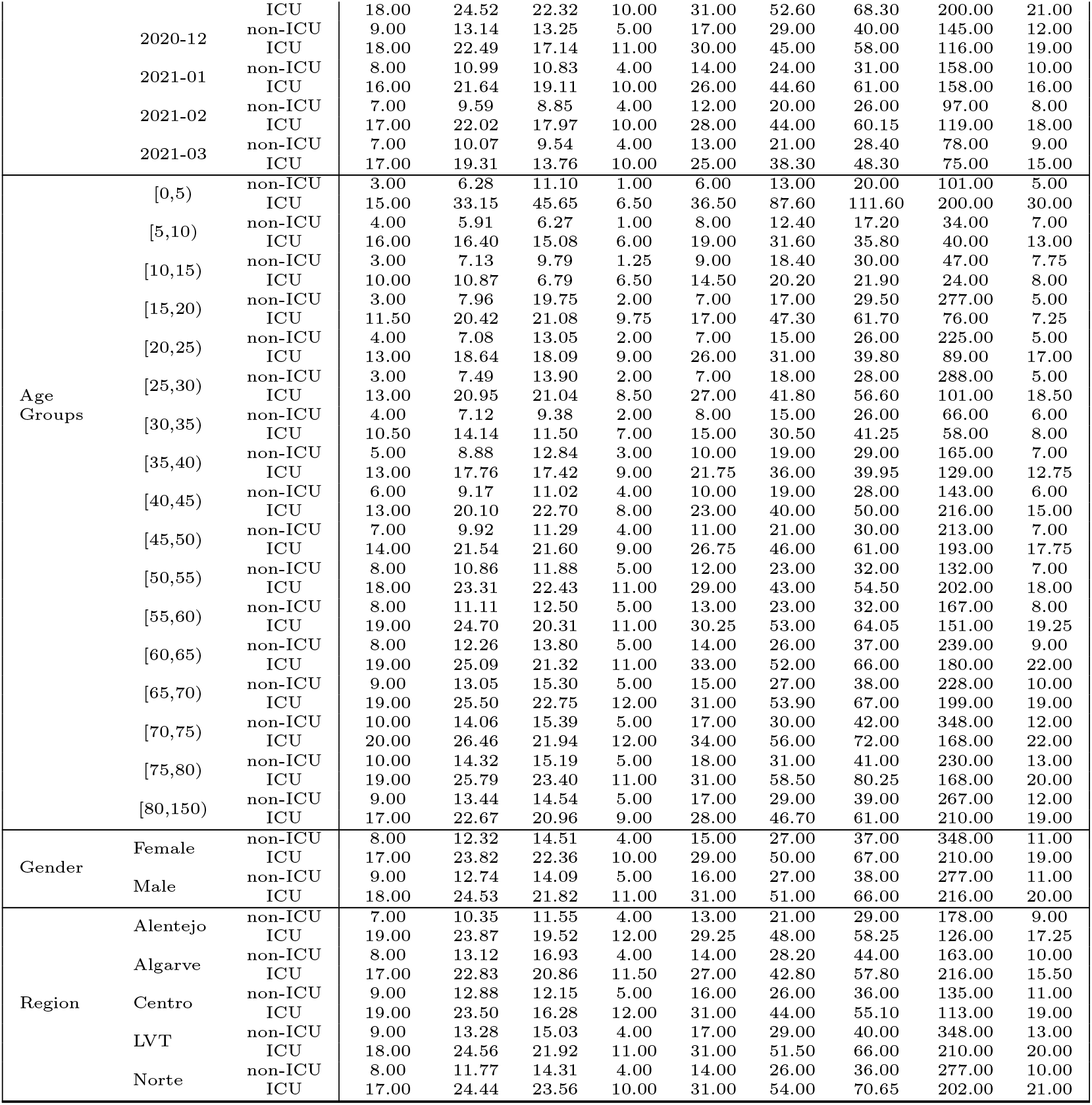
Summary statistics of the LoS, by type of hospitalisation (non-ICU and ICU), entry month, age group, gender and region.

**Table 13:** Summary statistics of the TuD, by type of hospitalisation (non-ICU and ICU), month of discharge, age group, gender and region.

| Variable | Group | Local | median | mean | sd | 25% | 75% | 90% | 95% | Max | IQR |
| --- | --- | --- | --- | --- | --- | --- | --- | --- | --- | --- | --- |
| Month | 2020-03 | non-ICU | 3.00 | 4.59 | 4.22 | 2.00 | 6.00 | 9.00 | 11.40 | 24.00 | 4.00 |
|  |  | ICU | 6.50 | 7.89 | 6.61 | 2.00 | 11.25 | 16.60 | 19.30 | 26.00 | 9.25 |
|  | 2020-04 | non-ICU | 7.00 | 9.43 | 8.32 | 3.00 | 13.00 | 21.00 | 25.30 | 49.00 | 10.00 |
|  |  | ICU | 12.00 | 14.24 | 9.83 | 7.00 | 22.00 | 29.80 | 32.90 | 38.00 | 15.00 |
|  | 2020-05 | non-ICU | 12.00 | 18.06 | 16.78 | 6.00 | 25.25 | 46.00 | 52.00 | 75.00 | 19.25 |
|  |  | ICU | 24.50 | 28.77 | 18.05 | 18.75 | 35.25 | 49.70 | 62.40 | 80.00 | 16.50 |
|  | 2020-06 | non-ICU | 8.00 | 15.22 | 18.08 | 5.00 | 18.00 | 38.20 | 50.10 | 98.00 | 13.00 |
|  |  | ICU | 26.00 | 32.21 | 29.02 | 10.00 | 43.00 | 75.60 | 91.00 | 101.00 | 33.00 |
|  | 2020-07 | non-ICU | 8.00 | 15.46 | 19.48 | 4.50 | 18.50 | 33.20 | 44.10 | 135.00 | 14.00 |
|  |  | ICU | 12.00 | 24.36 | 32.26 | 6.00 | 24.75 | 61.40 | 110.20 | 139.00 | 18.75 |
|  | 2020-08 | non-ICU | 11.50 | 17.62 | 17.88 | 5.00 | 22.50 | 39.50 | 61.00 | 90.00 | 17.50 |
|  |  | ICU | 15.50 | 28.06 | 35.07 | 9.25 | 29.00 | 56.50 | 73.65 | 151.00 | 19.75 |
|  | 2020-09 | non-ICU | 8.00 | 11.34 | 12.33 | 4.00 | 14.00 | 21.30 | 37.10 | 85.00 | 10.00 |
|  |  | ICU | 12.00 | 13.79 | 9.15 | 9.00 | 19.00 | 22.20 | 29.40 | 43.00 | 10.00 |
|  | 2020-10 | non-ICU | 7.00 | 10.45 | 11.91 | 3.00 | 13.00 | 22.00 | 31.60 | 90.00 | 10.00 |
|  |  | ICU | 13.00 | 17.87 | 18.22 | 6.50 | 24.00 | 37.60 | 46.00 | 141.00 | 17.50 |
|  | 2020-11 | non-ICU | 6.00 | 9.36 | 11.60 | 3.00 | 12.00 | 19.00 | 26.55 | 163.00 | 9.00 |
|  |  | ICU | 13.00 | 15.89 | 13.61 | 7.00 | 21.00 | 31.00 | 39.35 | 92.00 | 14.00 |
|  | 2020-12 | non-ICU | 8.00 | 11.03 | 12.37 | 4.00 | 14.00 | 22.90 | 30.45 | 206.00 | 10.00 |
|  |  | ICU | 17.00 | 19.01 | 12.10 | 10.00 | 26.25 | 35.00 | 40.25 | 82.00 | 16.25 |
|  | 2021-01 | non-ICU | 6.50 | 10.20 | 11.91 | 3.00 | 13.00 | 23.00 | 32.00 | 149.00 | 10.00 |
|  |  | ICU | 13.00 | 17.46 | 15.79 | 8.00 | 21.75 | 37.00 | 51.75 | 98.00 | 13.75 |
|  | 2021-02 | non-ICU | 8.00 | 10.87 | 10.43 | 4.00 | 14.00 | 24.00 | 31.00 | 106.00 | 10.00 |
|  |  | ICU | 17.00 | 20.44 | 17.46 | 12.00 | 25.00 | 32.40 | 45.70 | 180.00 | 13.00 |
|  | 2021-03 | non-ICU | 14.00 | 19.73 | 20.59 | 7.00 | 25.00 | 41.00 | 58.20 | 178.00 | 18.00 |
|  |  | ICU | 26.00 | 27.50 | 17.49 | 17.00 | 36.00 | 49.00 | 56.00 | 134.00 | 19.00 |
|  | 2021-04 | non-ICU | 31.00 | 37.34 | 30.14 | 17.00 | 43.50 | 80.40 | 84.80 | 142.00 | 26.50 |
|  |  | ICU | 35.00 | 39.84 | 24.17 | 20.00 | 64.00 | 75.60 | 84.20 | 89.00 | 44.00 |
|  | 2021-05 | non-ICU | 96.50 | 114.75 | 86.61 | 52.25 | 159.00 | 200.40 | 214.20 | 228.00 | 106.75 |
|  |  | ICU | 73.00 | 84.70 | 49.85 | 46.25 | 105.75 | 128.80 | 163.90 | 199.00 | 59.50 |
| Age Groups | [0,5) | non-ICU | 2.00 | 2.00 | - | 2.00 | 2.00 | 2.00 | 2.00 | 2.00 | 0.00 |
|  |  | ICU | 18.00 | 18.00 | - | 18.00 | 18.00 | 18.00 | 18.00 | 18.00 | 0.00 |
|  | [5,10) | non-ICU | - | - | - | - | - | - | - | - | - |
|  |  | ICU | - | - | - | - | - | - | - | - | - |
|  | [10,15) | non-ICU | - | - | - | - | - | - | - | - | - |
|  |  | ICU | 1.00 | 1.00 | - | 1.00 | 1.00 | 1.00 | 1.00 | 1.00 | 0.00 |
|  | [15,20) | non-ICU | 48.00 | 48.00 | - | 48.00 | 48.00 | 48.00 | 48.00 | 48.00 | 0.00 |
|  |  | ICU | - | - | - | - | - | - | - | - | - |
|  | [20,25) | non-ICU | 23.00 | 23.00 | - | 23.00 | 23.00 | 23.00 | 23.00 | 23.00 | 0.00 |
|  |  | ICU | - | - | - | - | - | - | - | - | - |
|  | [25,30) | non-ICU | 6.00 | 14.20 | 23.48 | 2.00 | 6.00 | 36.00 | 46.00 | 56.00 | 4.00 |
|  |  | ICU | 30.00 | 30.00 | - | 30.00 | 30.00 | 30.00 | 30.00 | 30.00 | 0.00 |
|  | [30,35) | non-ICU | 6.50 | 10.75 | 11.66 | 3.50 | 13.50 | 21.60 | 29.30 | 37.00 | 10.00 |
|  |  | ICU | 7.00 | 8.29 | 4.27 | 6.00 | 11.50 | 13.40 | 13.70 | 14.00 | 5.50 |
|  | [35,40) | non-ICU | 6.00 | 15.59 | 23.32 | 5.00 | 17.00 | 29.80 | 49.20 | 98.00 | 12.00 |
|  |  | ICU | 17.00 | 24.83 | 32.34 | 7.50 | 23.50 | 40.30 | 77.55 | 121.00 | 16.00 |
|  | [40,45) | non-ICU | 8.00 | 15.50 | 14.80 | 4.75 | 26.75 | 39.00 | 40.95 | 51.00 | 22.00 |
|  |  | ICU | 11.50 | 20.75 | 27.95 | 7.25 | 23.50 | 37.40 | 46.10 | 139.00 | 16.25 |
|  | [45,50) | non-ICU | 9.50 | 14.62 | 15.81 | 3.75 | 21.00 | 29.30 | 45.45 | 83.00 | 17.25 |
|  |  | ICU | 16.00 | 26.24 | 29.27 | 8.00 | 30.00 | 61.80 | 78.60 | 142.00 | 22.00 |
|  | [50,55) | non-ICU | 10.00 | 16.17 | 16.96 | 5.00 | 22.50 | 37.70 | 51.45 | 83.00 | 17.50 |
|  |  | ICU | 20.00 | 24.91 | 22.40 | 12.00 | 30.75 | 46.00 | 60.70 | 141.00 | 18.75 |
|  | [55,60) | non-ICU | 11.00 | 16.78 | 19.55 | 5.00 | 21.75 | 37.00 | 51.95 | 142.00 | 16.75 |
|  |  | ICU | 19.00 | 24.48 | 22.47 | 11.00 | 29.00 | 44.40 | 62.70 | 151.00 | 18.00 |
|  | [60,65) | non-ICU | 8.00 | 13.88 | 17.73 | 3.50 | 18.50 | 32.00 | 43.00 | 178.00 | 15.00 |
|  |  | ICU | 17.00 | 21.56 | 19.59 | 10.00 | 27.00 | 43.60 | 56.95 | 180.00 | 17.00 |
|  | [65,70) | non-ICU | 9.00 | 13.79 | 14.88 | 4.00 | 19.00 | 31.00 | 43.80 | 99.00 | 15.00 |
|  |  | ICU | 16.00 | 21.21 | 20.63 | 9.00 | 27.00 | 41.00 | 55.00 | 199.00 | 18.00 |
|  | [70,75) | non-ICU | 8.00 | 13.58 | 15.70 | 4.00 | 17.00 | 30.70 | 41.00 | 132.00 | 13.00 |
|  |  | ICU | 16.00 | 19.24 | 14.58 | 10.00 | 26.00 | 35.20 | 45.00 | 101.00 | 16.00 |
|  | [75,80) | non-ICU | 8.00 | 11.53 | 12.21 | 4.00 | 15.00 | 25.00 | 33.45 | 149.00 | 11.00 |
|  |  | ICU | 16.00 | 19.20 | 16.38 | 9.00 | 24.00 | 36.00 | 49.00 | 112.00 | 15.00 |
|  | [80,150) | non-ICU | 7.00 | 10.09 | 11.68 | 3.00 | 13.00 | 22.00 | 29.00 | 228.00 | 10.00 |
|  |  | ICU | 12.00 | 15.77 | 14.07 | 7.00 | 20.00 | 32.00 | 43.00 | 115.00 | 13.00 |
| Gender | Female | non-ICU | 7.00 | 10.22 | 12.30 | 3.00 | 13.00 | 22.00 | 31.00 | 206.00 | 10.00 |
|  |  | ICU | 15.00 | 18.99 | 18.87 | 8.00 | 24.00 | 37.00 | 53.00 | 180.00 | 16.00 |
|  | Male | non-ICU | 8.00 | 11.74 | 13.19 | 4.00 | 15.00 | 25.00 | 34.00 | 228.00 | 11.00 |
|  |  | ICU | 17.00 | 20.47 | 18.27 | 9.00 | 26.00 | 39.00 | 53.00 | 199.00 | 17.00 |
| Region | Alentejo | non-ICU | 5.00 | 7.83 | 10.97 | 3.00 | 10.00 | 16.00 | 22.25 | 178.00 | 7.00 |
|  |  | ICU | 17.00 | 20.84 | 18.29 | 7.00 | 23.50 | 51.50 | 58.75 | 89.00 | 16.50 |
|  | Algarve | non-ICU | 8.00 | 13.94 | 20.79 | 4.00 | 15.00 | 28.80 | 41.80 | 163.00 | 11.00 |
|  |  | ICU | 16.00 | 18.92 | 11.30 | 11.25 | 26.00 | 30.00 | 38.75 | 64.00 | 14.75 |
|  | Centro | non-ICU | 7.00 | 10.51 | 10.97 | 4.00 | 13.00 | 23.00 | 29.00 | 135.00 | 9.00 |
|  |  | ICU | 16.00 | 18.50 | 13.15 | 10.00 | 24.25 | 34.00 | 40.45 | 96.00 | 14.25 |
|  | LVT | non-ICU | 8.00 | 12.08 | 13.94 | 4.00 | 16.00 | 26.00 | 36.00 | 228.00 | 12.00 |
|  |  | ICU | 15.00 | 19.56 | 17.76 | 8.00 | 26.00 | 38.60 | 54.60 | 180.00 | 18.00 |
|  | Norte | non-ICU | 7.00 | 10.29 | 11.59 | 3.00 | 13.00 | 22.00 | 32.00 | 163.00 | 10.00 |
|  |  | ICU | 16.00 | 21.19 | 21.40 | 8.00 | 27.00 | 43.00 | 59.50 | 199.00 | 19.00 |

#### 3.1.2. Characterization by age

It is well known that COVID-19 affects more the elderly, increasing the risk of hospitalisation, severe illness and death. This was visible in the data as fatality rate increases with age, as shown in figure 5(b), as well as a higher percentage of hospitalized population (appendix A.5, figure 17(a) and (b)). The probability of being admitted to ICU was also higher for patients with 60 to 69 years of age, decreasing afterwards. This makes us conclude that the probability of entering ICU for older patients is lower.

**Figure 16:**
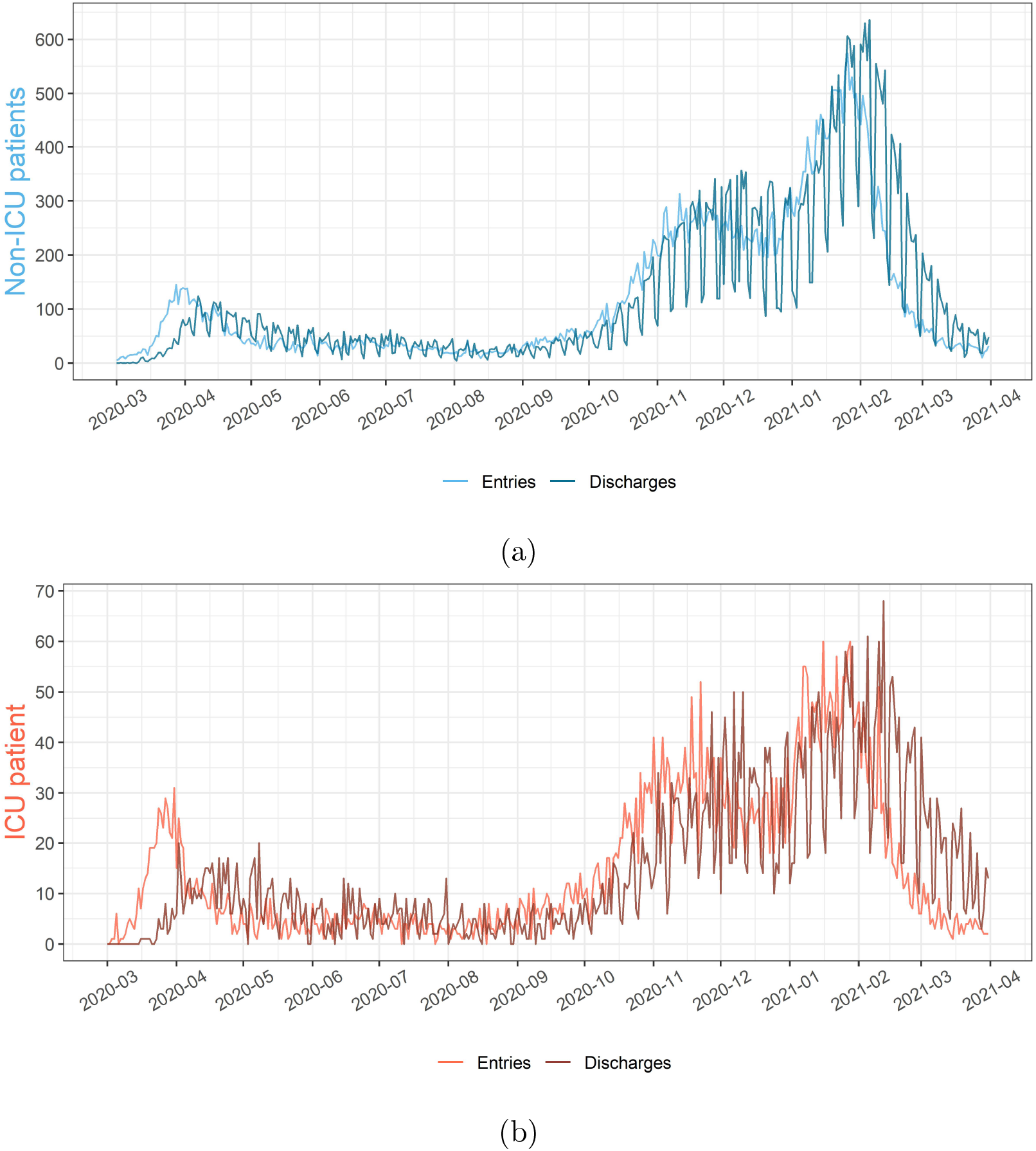
Comparing entries and exits by time, for (a)non-ICU and (b)ICU patients.

**Figure 17:**
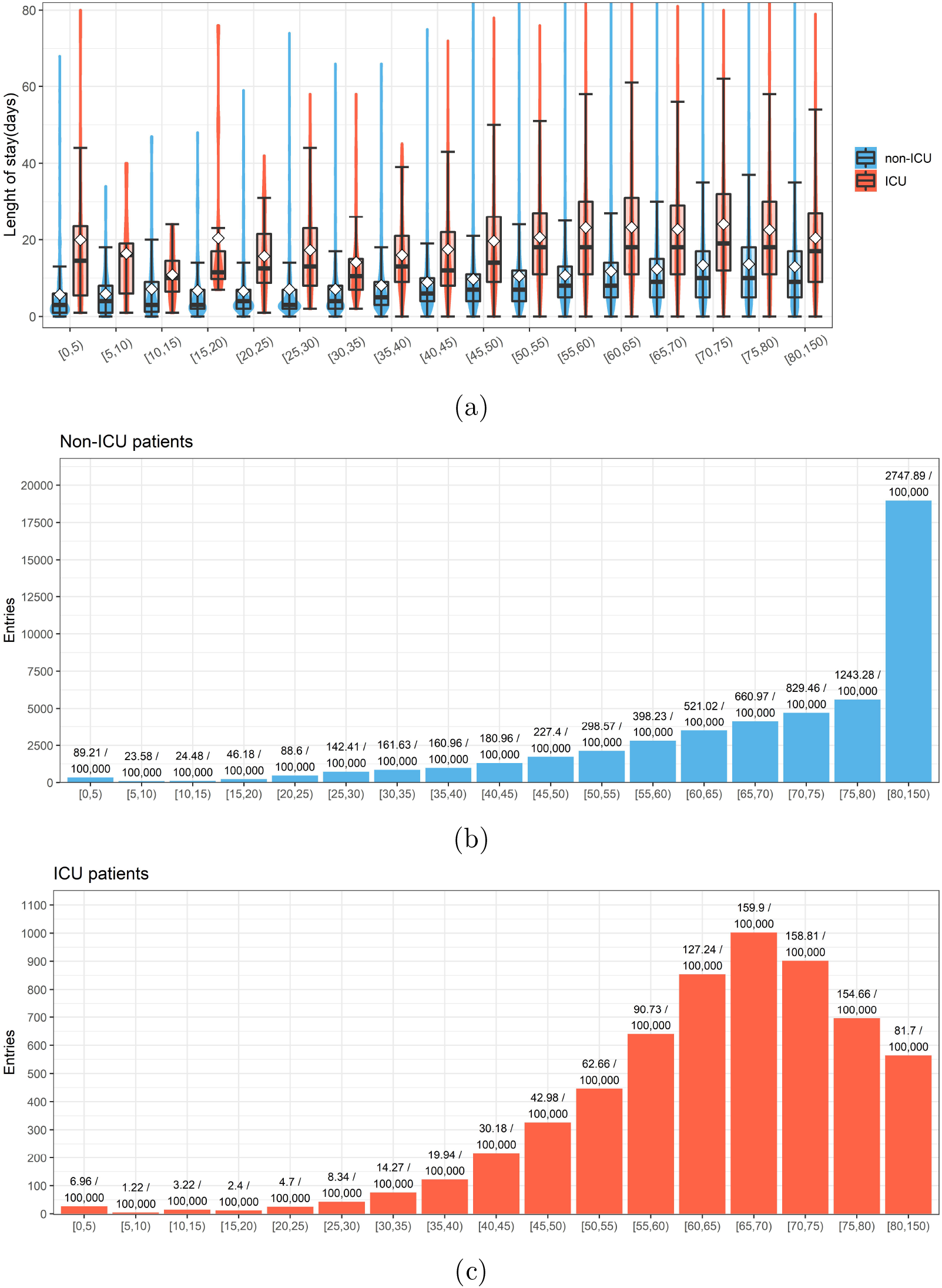
Characterization of the age groups. **(a)**Violin/box plot of the LoS by age groups (white diamonds represent the mean). Histograms of the number of entries, by age group, for **(b)**non-ICU and **(c)**ICU patients

**Figure 18:**
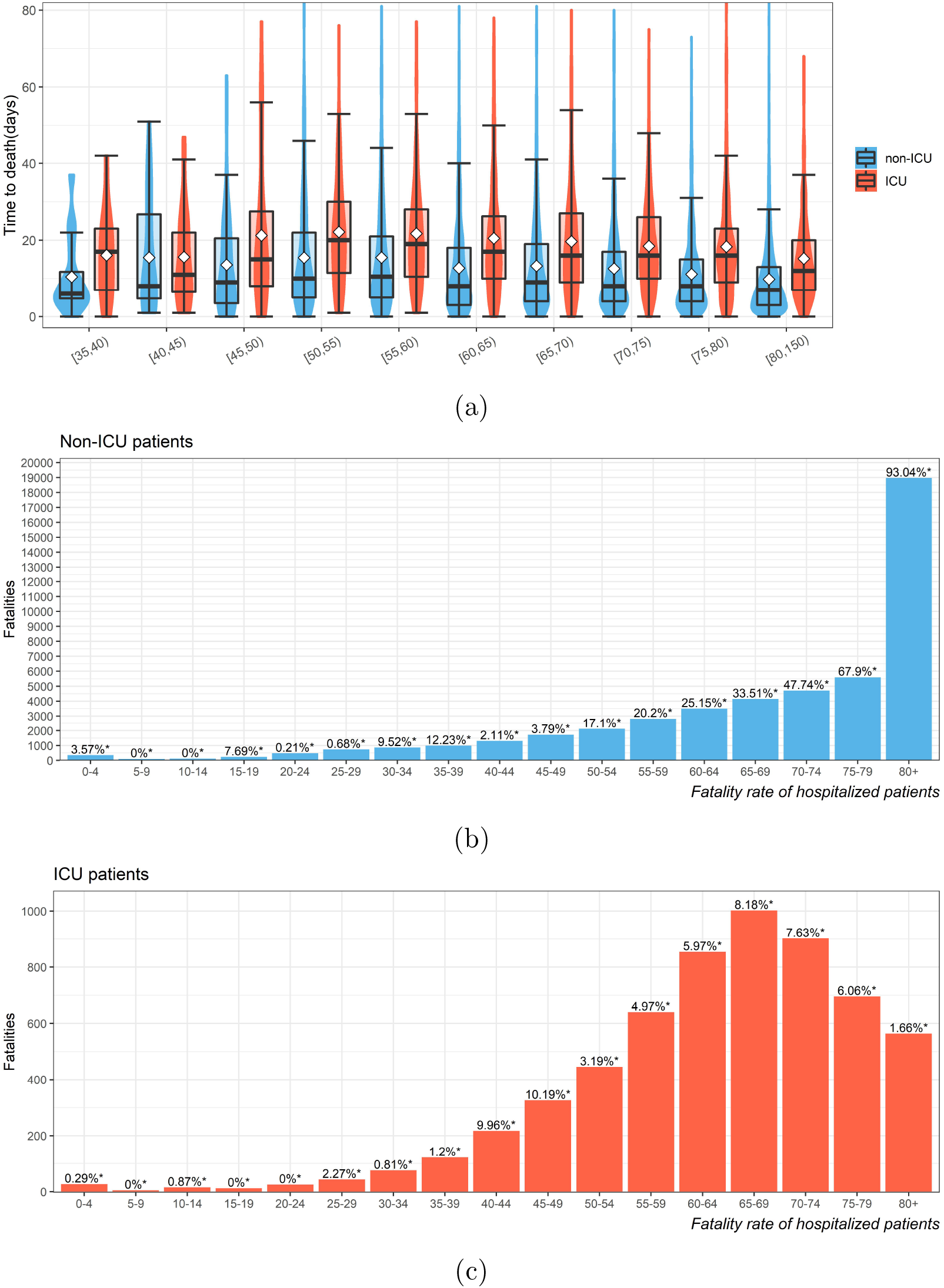
**(a)**Violin/box plot of the TuD by age groups (white diamonds represent the mean). Histogram of the fatalities, by age group, of **(b)**non-ICU and **(c)**ICU patients

The group from 10 to 15 years old is the one with the lower number of entries, but where the percentage of ICU patients and the fatality rate is higher than the 0 to 10 groups and the 15 to 20 group. This anomaly is not exclusive to Portugal and was reported by the Center of Disease Control and Prevention (CDC)[10].

#### 3.1.3. Characterization by gender

Looking at gender differences in COVID-19 hospitalisation, there is a clear difference in the total number hospitalisation, where males were more hospitalized than females, with percentage of the population hospitalized in non-ICU of 0.51% for males and 0.45% for females, and for ICU 0.08% for males and 0.03% for females. Male patients also have a higher fatality rate than females. For non-ICU, males patients have a 23.23% mortality rate and female patients have 21.95%, for ICU patients males have 33.81% and females 30.70 %.

Violin/box plots for the LoS considering gender and age groups are presented in appendix A.5 figure 20, where it is observable that the LoS are similar between group’s gender, being the only exceptions the groups of the 25-30 and 30-35 for non-ICU patients were a large difference between genders arises. In the case of TuD there is also no observable difference, as seen in appendix A.5 figure 19.

**Figure 19:**
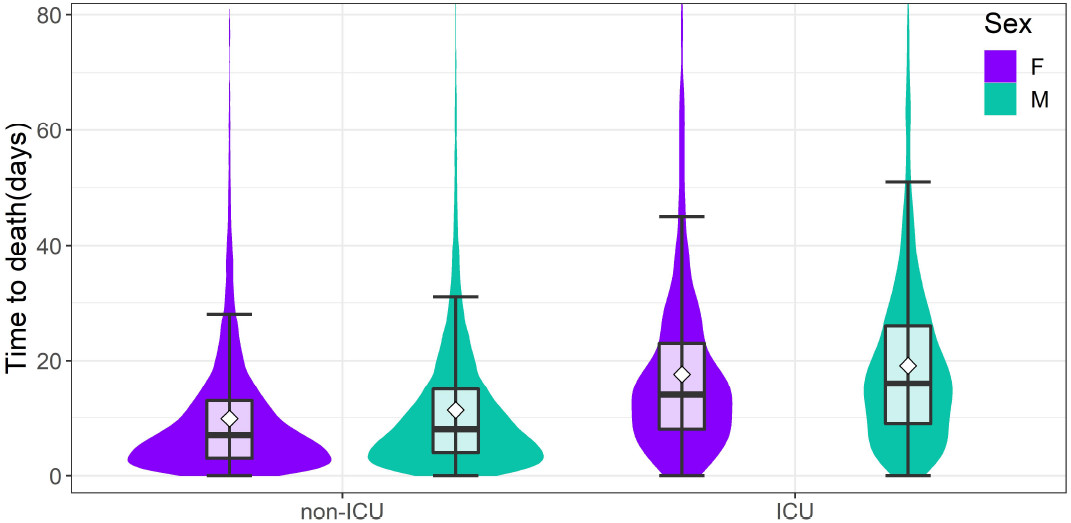
Violin/box plot of the TuD by gender and type of hospitalisation (white diamonds represent the mean).

**Figure 20:**
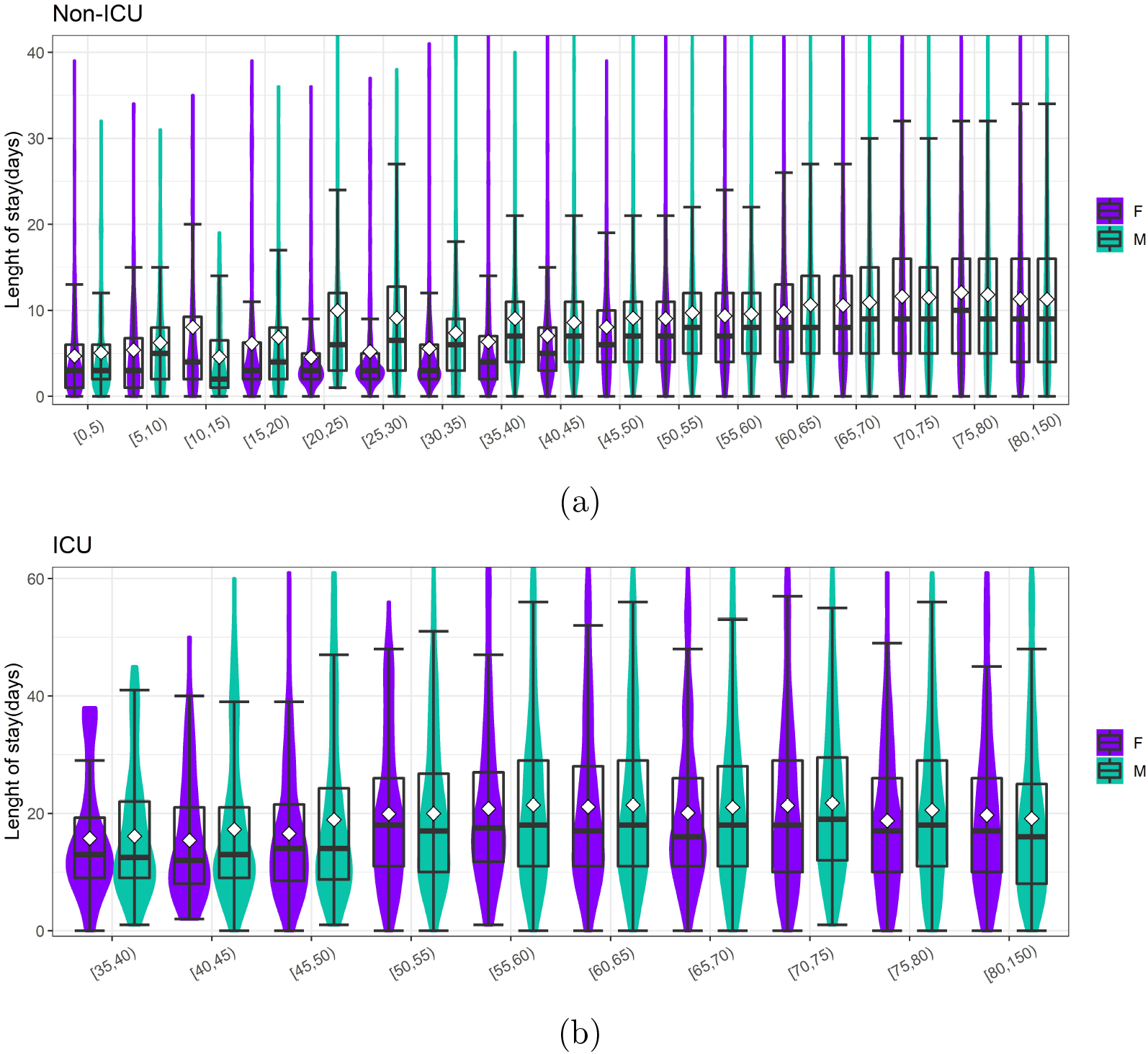
Violin/box plot of the LoS by gender and type of hospitalisation (white diamonds represent the mean).

**Figure 21:**
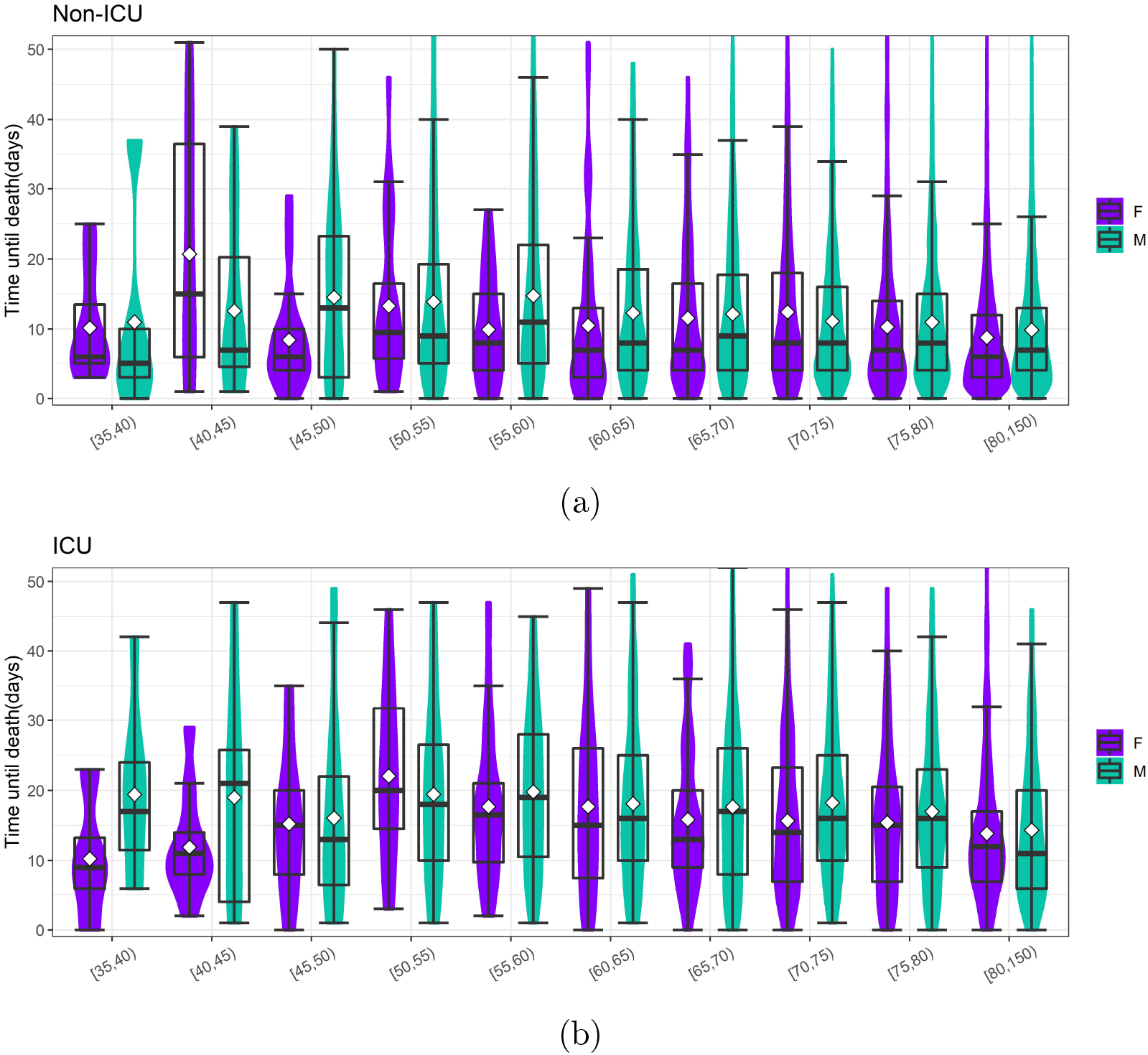
Violin/box plot of the TuD by gender and type of hospitalisation (white diamonds represent the mean).

In the Appendix A.5 figure 22 multiple graphs considering the differences between gender and type of hospitalization are presented. It is visible a lower number of entries and fatalities in females in general, with the females only being the group with more entries and fatalities in the 80 and over group for non-ICU patient type. Males have specially high numbers of entries and fatalities in ICU when compared to females.

**Figure 22:**
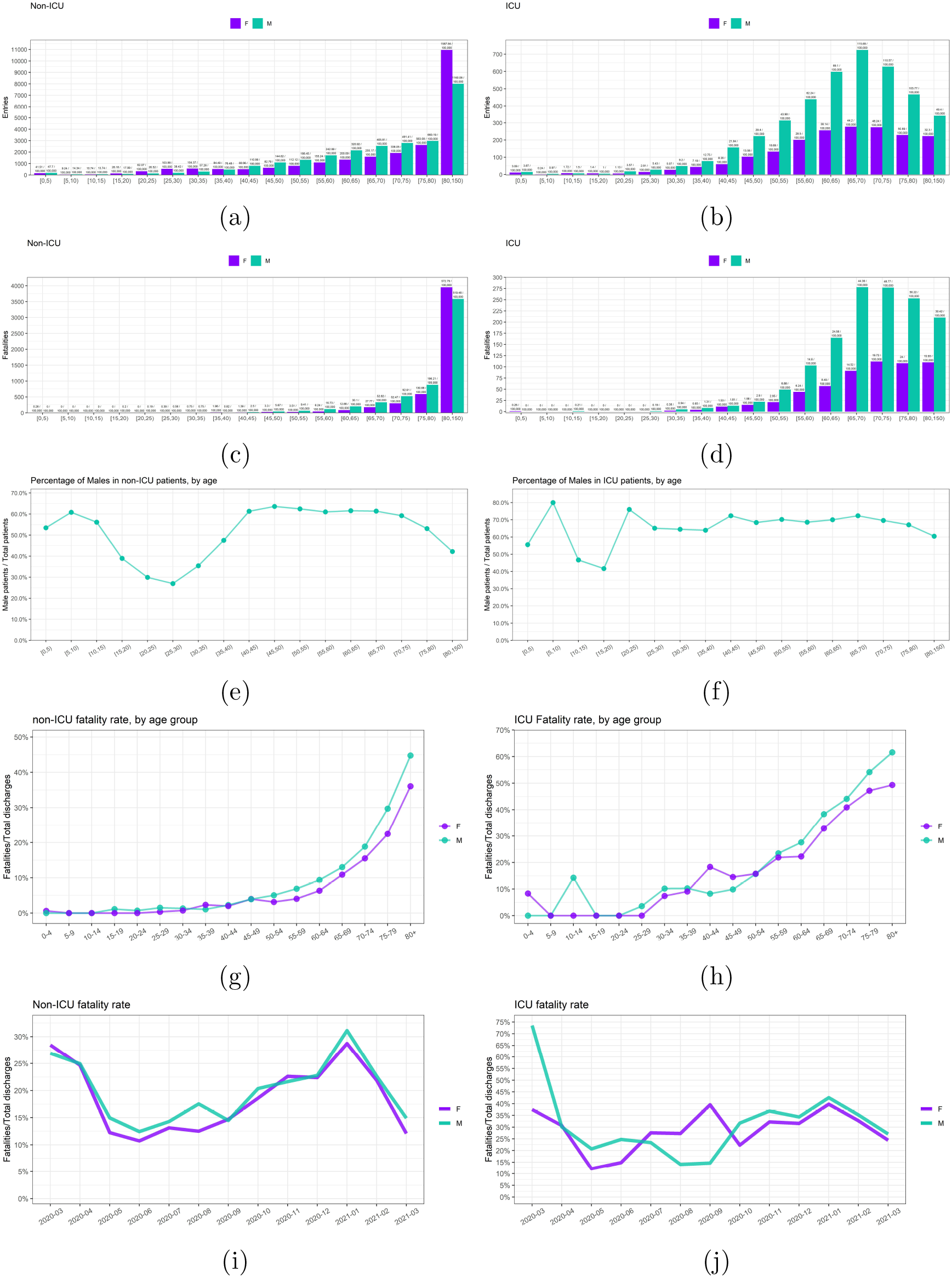
(a,b,c,d)Histograms with entries and fatalities for ICU and non-ICU patients. Linegraphs of fatalities rates by group for (e)non-ICU and (f)ICU patients. Percentage of males in (g)non-ICU and (h)ICU patients, for age group. Fatality rate over time for (i)non-ICU and (j)ICU.

**Figure 23:**
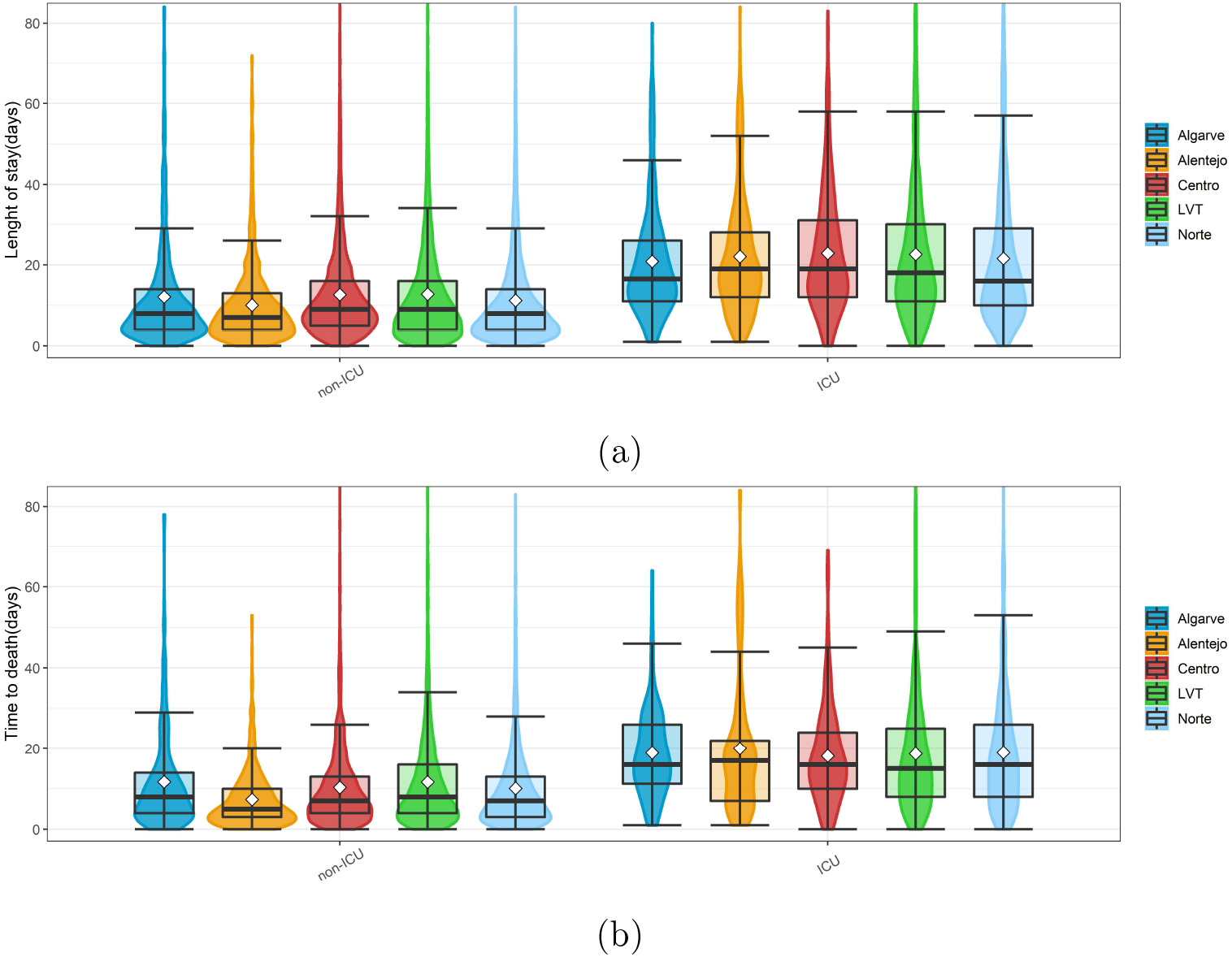
Violin/box plots of (a)LoS and (b)TuD by region (white diamonds represent the mean).

#### 3.1.4. Characterization by region

The SARS-CoV2 geographical spread through Portugal was not uniform, with regions having increases and decreases in incidence in different periods, although the increase in cases in one region seems to spread to all other regions, as can be seen in appendix A.5 figure 25.

**Figure 24:**
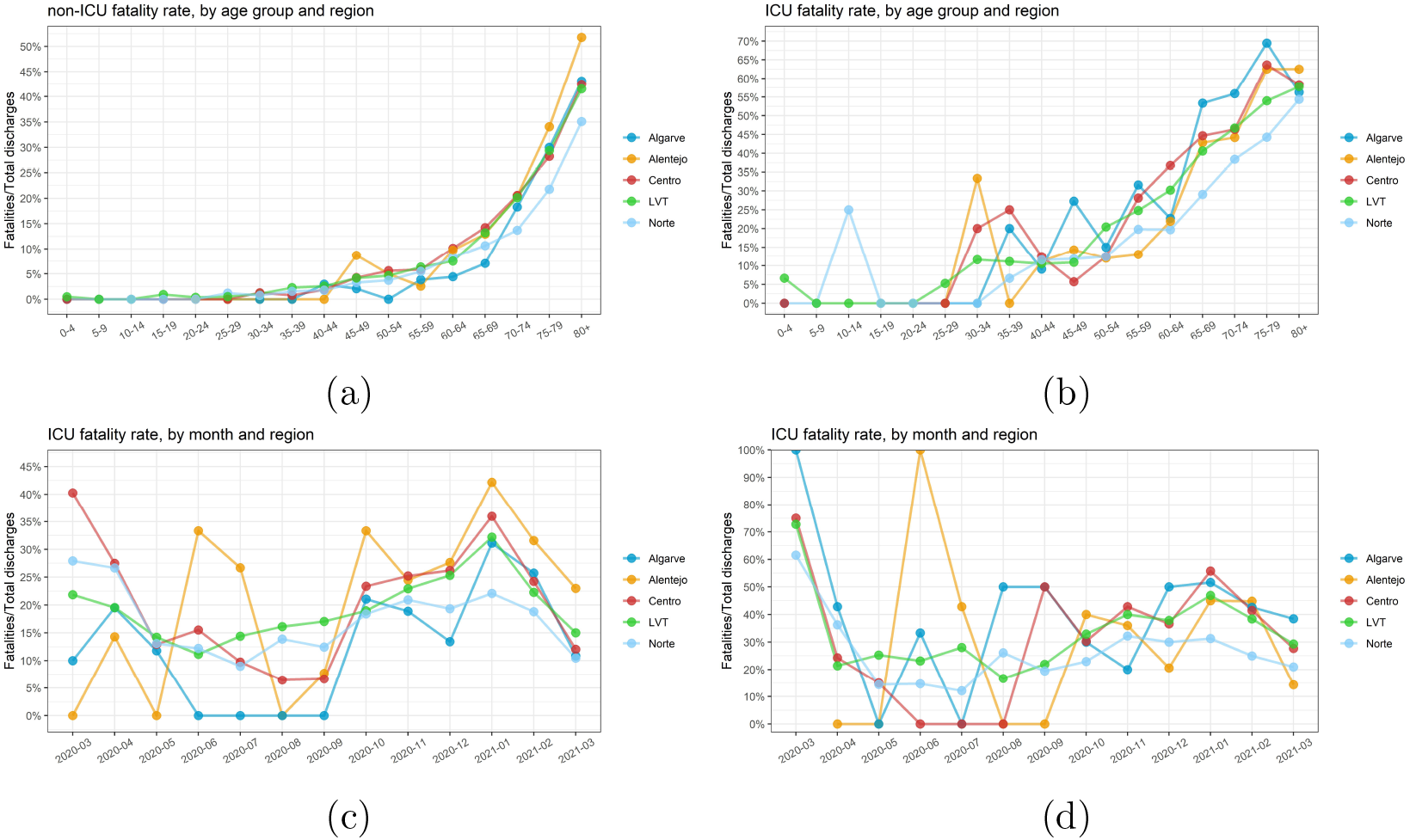
Linegraphs of fatalities rates by age group and region for (a) non-ICU(a) and (b)ICU patients. Fatality rate by region and over time for (c)non-ICU and (d)ICU (white diamonds represent the mean).

**Figure 25:**
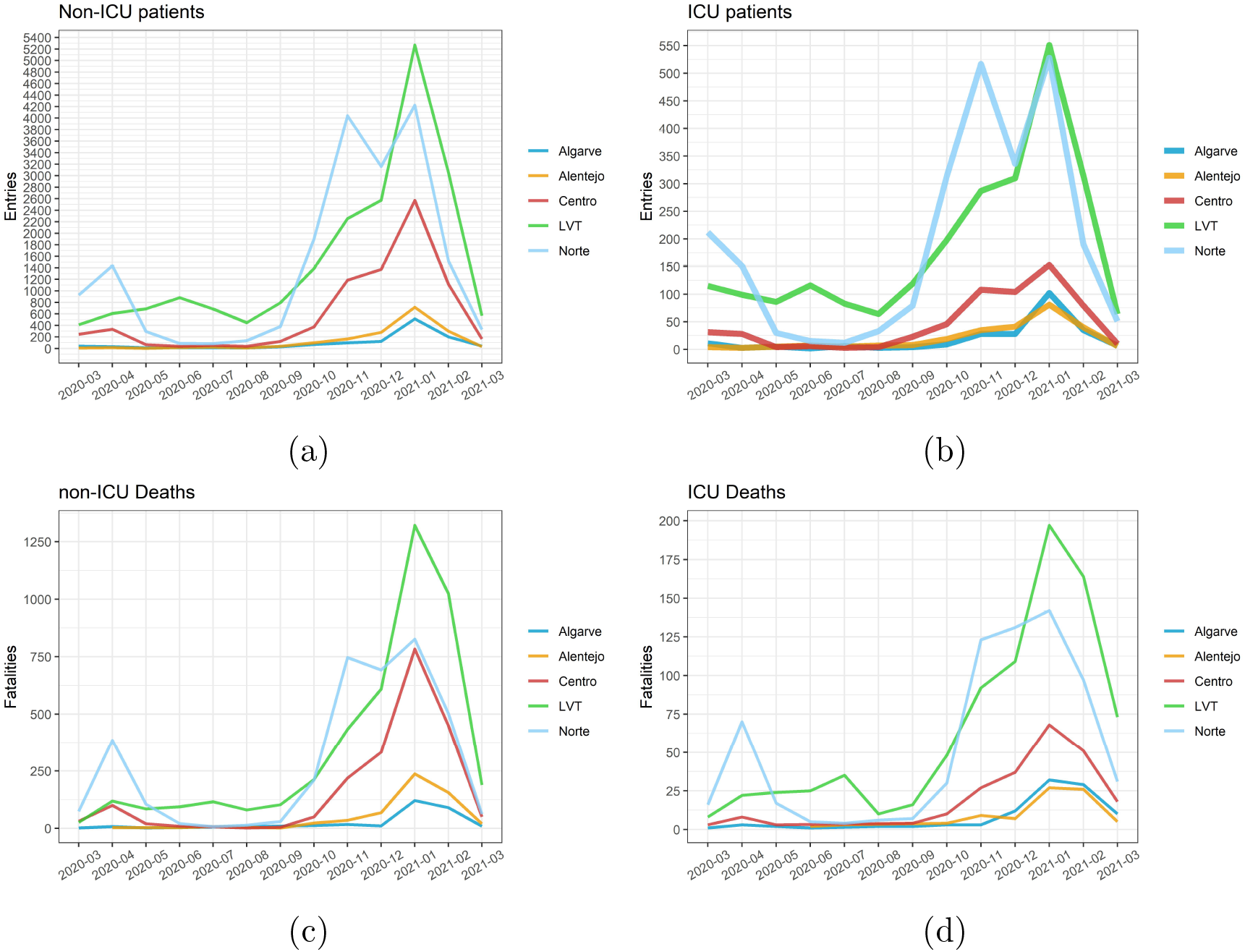
Linegraphs of entries by region for (a)non-ICU and (b)ICU patients entries, and fatalities for (c)non-ICU and (d)ICU patients fatalities

This leads to the differences in the percentage of the population admitted to hospital by region, observed in figure 8, with the *LVT* region being the one with the biggest percentage of hospitalization rate, followed by the *Norte* and *Centro* regions, other very populous regions. This could be explained by the higher population density of these regions that makes social distancing less efficient and by the transfer of patients from smaller hospitals located in less populated regions to larger ones, in big cities like Lisboa (*LVT* region), Porto (*Norte* region) and Coimbra (*Centro*).

No major difference was found between LoS and TuD by region, as seen in appendix A.5 figure 25. Looking at the percentage of patients requiring ICU treatment the *Centro* region has the lower and the *Algarve* region the higher percentage.

### 3.2. Distribution fitting

An analysis of the distribution of the LoS was conducted as explained in section 2. Using the Cullen and Frey graphs (figure 11) we chose the distributions that best fitted each sample of the LoS type of hospitalisation, non-ICU and ICU. From the interpretations of those graphs the gamma, lognormal and Weibull distributions were the ones chosen to be fitted to the data. A table with the summary statistics used to plot these graphs is available in the appendix A.6 table 14.

**Table 14:** LoS summary statistics for the Cullen and Frey graph (Figure 11).

| Local | Hosp | ICU |
| --- | --- | --- |
| min | 1 | 1 |
| max | 348 | 216 |
| median | 9 | 18 |
| mean | 12.81596 | 24.43704 |
| estimated sd | 14.33082 | 21.97512 |
| estimated skewness | 4.475019 | 2.608978 |
| estimated kurtosis | 45.76527 | 13.74304 |

After fitting the previously chosen distributions, for each type of hospitalisation (non-ICU and ICU), using the maximum likelihood estimation method, we compared them by using goodness-of-fit plots and the goodness of fit statistics and criteria supplied in the *fitdistrplus* package. We also estimated the gamma and log-normal distribution by fitting using the Left-tail AD variant of the Anderson-Darling distance. Tables with all the results are present in appendix A.6 (see 16 and 17).

The graphs in figure 12 represent all fitted theoretical distributions to the data for non-ICU and ICU patients. All estimated parameters are available in the appendix A.6 in table 15. For non-ICU patients the lognormal distribution (mean-log 2.1195 and sd-log 0.9496) was the distribution with the lower goodness-of-fit(GoF) statistics, except for the Anderson-Darling statistic that was lower in the lognormal ADL distribution (mean-log 2.1465 and sd-log 0.9886). The lognormal distribution (mean-log 2.8583 and sd-log 0.8655) also had lower GoF statistics for ICU patients, excepting for Kolmogorov-Smirnov and Anderson-Darling statistics that where lower for lognormal ADL distribution (mean-log 2.8986 and sd-log 0.8565).

**Table 15:** Estimated LoS distribution parameters.

| LoS |  | non-ICU | ICU |
| --- | --- | --- | --- |
| gamma | shape | 1.3009479 | 1.236824 |
|  | rate | 0.1015242 | 0.05061267 |
| log-normal | mean log | 2.119463 | 2.8582927 |
|  | sd log | 0.9495999 | 0.8655452 |
| Weibull | shape | 1.263677 | 1.253865 |
|  | scale | 13.09343 | 26.429118 |
| gamma ADL | shape | 1.5623940 | 1.236824 |
|  | rate | 0.1403541 | 0.05061267 |
| log-normal ADL | mean log | 2.1465055 | 2.8986155 |
|  | sd log | 0.9885536 | 0.8565485 |

The same analysis was carried out to the TuD, with the Cullen and Frey graphs and statistics being presented in figure 26 and table 18, and a table with the parameters for all fitted distribution in tables 19, in appendix A.6. Comparing goodness of fit statistics, for the non-ICU patients the lognormal distribution (mean-log 1.9684 and sd-log 0.9873) had lower values for Anderson-Darling statistic and Alkaike’s and Bayesian Information Criterion, with the gamma distribution (shape 1.2314 and rate 0.1088) havind lower values in the Kolmogorov-Smirnov and Cramer-von Mises statistics. For ICU patients the gamma distribution (shape 1.5799 and rate 0.0779) had lower values for all but Kolmogorov-Smirnov and Cramer-von Mises statistic that were lower for the gamma ADL distribution (rate 1.7360 and rate 0.0888).

**Figure 26:**
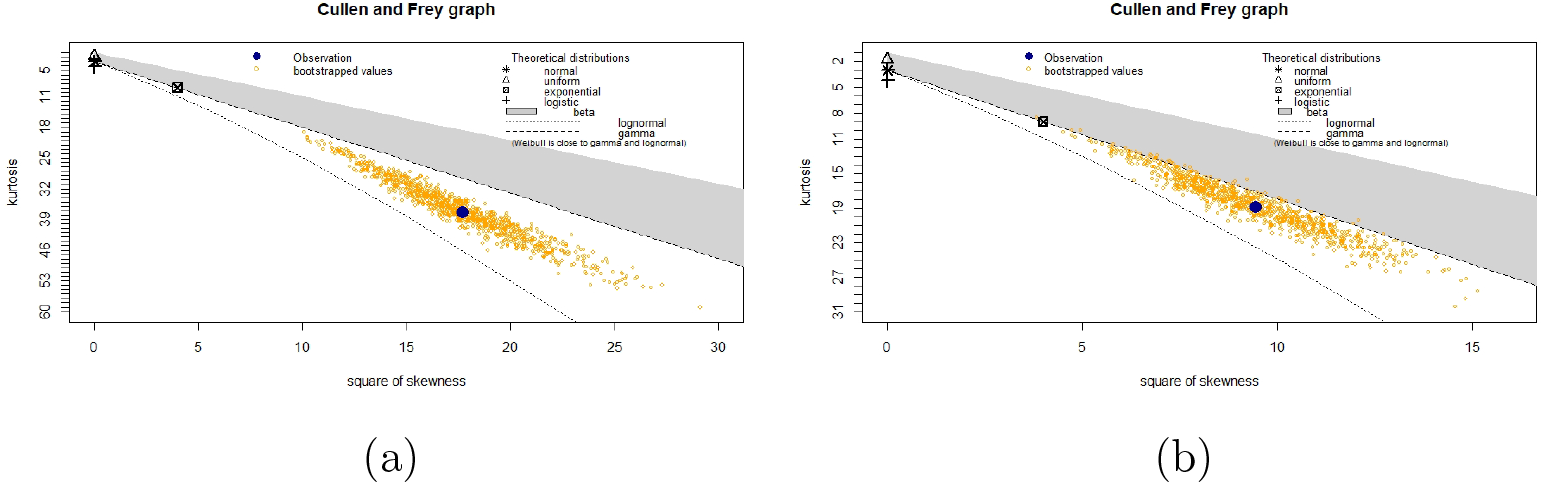
TuD Cullen and Frey graphs for (a)Non-ICU and (b)ICU patients.

**Table 16:** LoS goodness-of-fit statistics for non-ICU patients.

|  | gamma | lognormal | Weibull |
| --- | --- | --- | --- |
| Goodness-of-fit statistics |  |  |  |
| Kolmogorov-Smirnov statistic | 0.07854276 | 0.05344594 | 0.07335166 |
| Cramer-von Mises statistic | 63.00402211 | 15.96330257 | 66.41153408 |
| Anderson-Darling statistic | 365.04226391 | 127.87010687 | 446.79678352 |
| Goodness-of-fit criteria |  |  |  |
| Akaike's Information Criterion | 336511.5 | 332306.2 | 337756.5 |
| Bayesian Information Criterion | 336529.0 | 332323.8 | 337774.0 |
|  | gamma ADL | lognormal ADL |  |
| Goodness-of-fit statistics |  |  |  |
| Kolmogorov-Smirnov statistic | 0.04724813 | 0.04920051 |  |
| Cramer-von Mises statistic | 24.02657920 | 17.63190355 |  |
| Anderson-Darling statistic | Inf | 145.71971535 |  |
| Goodness-of-fit criteria |  |  |  |
| Akaike's Information Criterion | 339100.5 | 332491.9 |  |
| Bayesian Information Criterion | 339118.1 | 332509.4 |  |

**Table 17:** LoS goodness-of-fit statistics for ICU patients.

|  | gamma | lognormal | Weibull |
| --- | --- | --- | --- |
| Goodness-of-fit statistics |  |  |  |
| Kolmogorov-Smirnov statistic | 0.06457332 | 0.0566513 | 0.06895957 |
| Cramer-von Mises statistic | 2.8023735 | 1.76070045 | 8.34009006 |
| Anderson-Darling statistic | 19.9448961 | 12.46493868 | 50.56938674 |
| Goodness-of-fit criteria |  |  |  |
| Akaike's Information Criterion | 49117.16 | 49102.09 | 49350.59 |
| Bayesian Information Criterion | 49130.54 | 49115.47 | 49363.97 |
|  | gamma ADL | lognormal ADL |  |
| Goodness-of-fit statistics |  |  |  |
| Kolmogorov-Smirnov statistic | 0.04424102 | 0.03867471 |  |
| Cramer-von Mises statistic | 1.97564933 | 1.69481544 |  |
| Anderson-Darling statistic | 32.65844445 | 16.87600111 |  |
| Goodness-of-fit criteria |  |  |  |
| Akaike's Information Criterion | 49360.57 | 49116.56 |  |
| Bayesian Information Criterion | 49373.95 | 49129.94 |  |

**Table 18:**
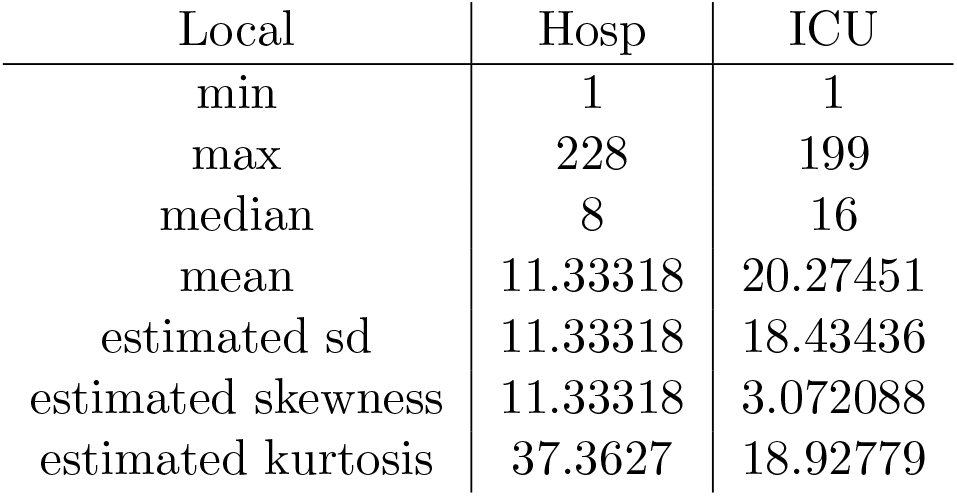
TuD Summary statistics for the Cullen and Frey graph (Figure 26).

**Table 19:** Estimated TuD distribution parameters.

| TuD |  | non-ICU | ICU |
| --- | --- | --- | --- |
| gamma | shape | 1.2314081 | 1.57990102 |
|  | rate | 0.1087638 | 0.07792103 |
| log-normal | meanlog | 1.9684210 | 2.660706 |
|  | sdlog | 0.9873343 | 0.898466 |
| Weibull | shape | 1.244128 | 1.246129 |
|  | scale | 21.859276 | 21.898314 |
| gamma ADL | shape | 1.73385831 | 1.73596577 |
|  | rate | 0.08887636 | 0.08882768 |
| log-normal ADL | meanlog | 2.734321 | 2.736472 |
|  | sdlog | 0.939225 | 0.938759 |

**Table 20:** TuD goodness-of-fit statistics for non-ICU patients.

|  | gamma | lognormal | Weibull |
| --- | --- | --- | --- |
| Goodness-of-fit statistics |  |  |  |
| Kolmogorov-Smirnov statistic | 0.06756849 | 0.05607134 | 0.07106338 |
| Cramer-von Mises statistic | 10.79585720 | 5.31072150 | 10.18207602 |
| Anderson-Darling statistic | 70.37506509 | 46.28643673 | 78.49461693 |
| Goodness-of-fit criteria |  |  |  |
| Akaike's Information Criterion | 73132.70 | 72288.43 | 73326.16 |
| Bayesian Information Criterion | 73147.26 | 72302.99 | 73340.71 |
|  | gamma ADL | lognormal ADL |  |
| Goodness-of-fit statistics |  |  |  |
| Kolmogorov-Smirnov statistic | 0.04833295 | 0.0518889 |  |
| Cramer-von Mises statistic | 4.80060941 | 5.4628413 |  |
| Anderson-Darling statistic | 66.41089597 | 52.02366989 |  |
| Goodness-of-fit criteria |  |  |  |
| Akaike's Information Criterion | 73448.81 | 72412.75 |  |
| Bayesian Information Criterion | 73463.37 | 72427.31 |  |

**Table 21:**
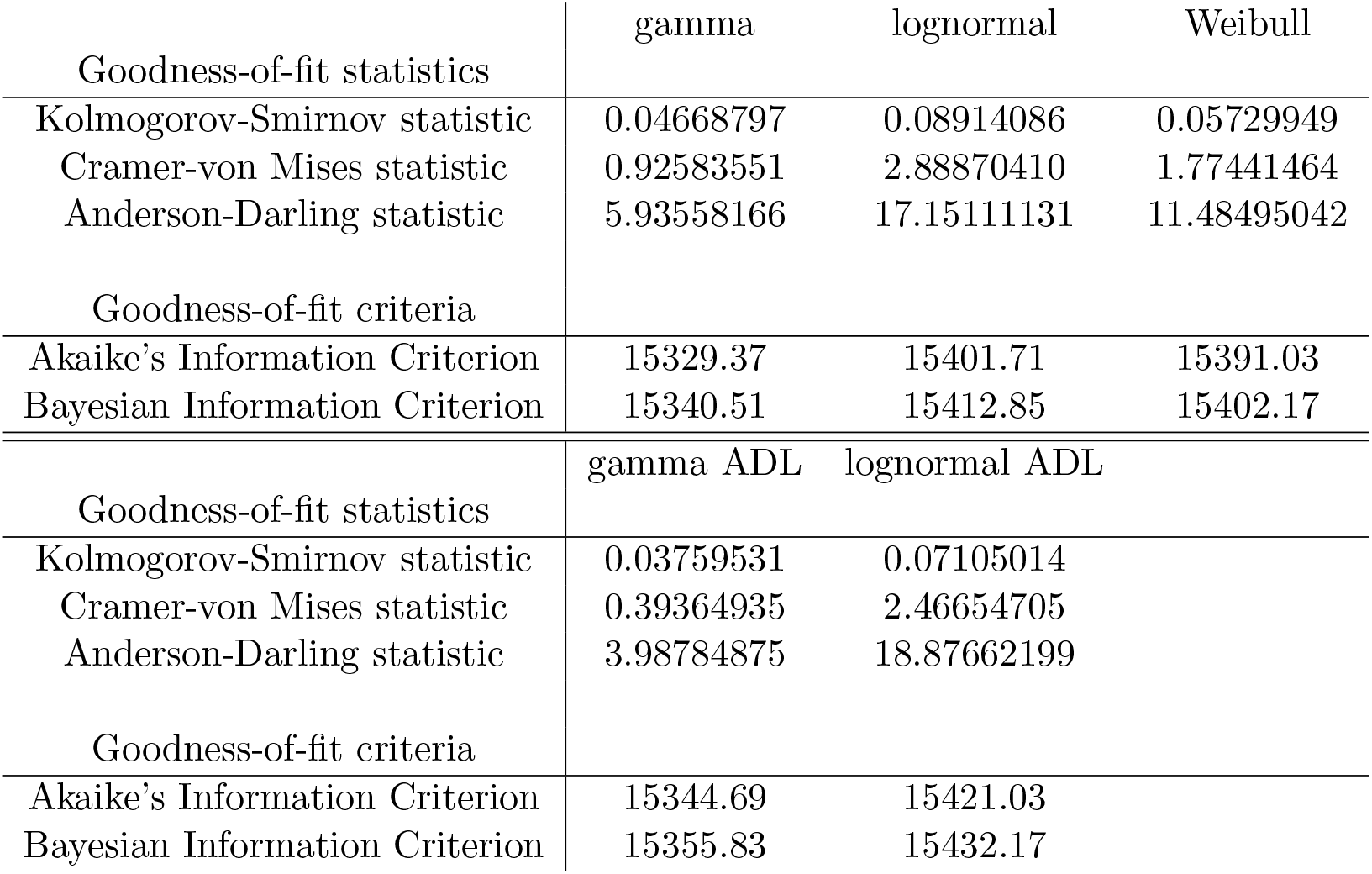
TuD goodness-of-fit statistics for ICU patients.

|  | gamma | lognormal | Weibull |
| --- | --- | --- | --- |
| Goodness-of-fit statistics |  |  |  |
| Kolmogorov-Smirnov statistic | 0.04668797 | 0.08914086 | 0.05729949 |
| Cramer-von Mises statistic | 0.92583551 | 2.88870410 | 1.77441464 |
| Anderson-Darling statistic | 5.93558166 | 17.15111131 | 11.48495042 |
| Goodness-of-fit criteria |  |  |  |
| Akaike's Information Criterion | 15329.37 | 15401.71 | 15391.03 |
| Bayesian Information Criterion | 15340.51 | 15412.85 | 15402.17 |
|  | gamma ADL | lognormal ADL |  |
| Goodness-of-fit statistics |  |  |  |
| Kolmogorov-Smirnov statistic | 0.03759531 | 0.07105014 |  |
| Cramer-von Mises statistic | 0.39364935 | 2.46654705 |  |
| Anderson-Darling statistic | 3.98784875 | 18.87662199 |  |
| Goodness-of-fit criteria |  |  |  |
| Akaike's Information Criterion | 15344.69 | 15421.03 |  |
| Bayesian Information Criterion | 15355.83 | 15432.17 |  |

## 4. DISCUSSION

From the analysis of Portuguese DRG data the mean length of stay (LoS) was estimated as 12.5 (IQR: 4-15) for patients without ICU hospitalization and 24.3 days (IQR: 11-30) for patients who received ICU treatments, with it being higher in the case of recovery when comparing with in the case of hospital fatality. No meaningful differences were found in LoS between genders and regions, but differences where found between age groups and over time, with older groups having a higher LoS. The mean time until death (TuD) and time until discharged (TuDisc) was also estimated as 11.01 and 12.94 for non-ICU patients and 20.06 and 26.39 days for ICU patients.

The percentage of patients that required entry in ICU was estimated as 10.92% and the fatality rate as 22.60% for non-ICU patients and 32.84% for ICU, with the percentage of ICU patients changing depending in patient age and the region of hospitalization. The fatality rate was different between age group, gender and region, for either non-ICU and ICU patients.

Various distributions were fitted to the density plots of the LoS and TuD, for either non-ICU and ICU patients, with the estimated distributions parameters being presented in the Appendix A.6 in table 15 for LoS and table 15 for TuD. Log-normal and log-normal ADL have better goodness-of-fit statistics for LoS of non-ICU and ICU and for TuD of non-ICU, gamma and gamma ADL have better fits for TuD of ICU patients.

In [9], the authors consider various distributions that were fitted to the hospitalized patients clinical database of Belgium, with the length of stay for patients that recover being estimated as between 5 days (in the young population) to 15.7 (in the elderly) and the median length of stay for patients that die between 5.7 days (in the elderly) and 12.2 days (in the working age population). Our study distinguishes between non-ICU and ICU patients but the LoS, TuD and TuDisc follow the trends presented by [9], increasing with the age of the patient. This study also refers that the LoS is higher for males, what was not observed in our study. A lognormal distribution was used to describe the LoS in hospital (log-mean = 2.480 and log-sd = 0.913; IBM = 76,865) and in ICU (log-mean = 2.183 and log-sd = 1.052; IBM = 7312), the LoS for recovery and death was also analysed.

From a retrospective cohort study of Portuguese surveillance data, Peixoto[16] identifies a consistent increase in risk of ICU admission with age until the 70-79 age group, with decreased risk for subsequent age groups. In our study the risk increases until age 60-75 age group, decreasing afterward, as can be seen in Figure 4(b). In regards to mortality, Peixoto[16] claims a constant increase of risk of death with age, which is also visible in our data, specially in in Figure 5(b).

In her paper Vekaria [13] estimates the various LoS, for the UK, using three different statistical methods. With LoS for non-ICU patients being between 8.4 and 9.3 days and for ICU being between 16.2 and 35.2 days. The study also presents the time from hospital entry to ICU as being between 2 to 4 days and the time from ICU exit to full hospital release being between 1.8 and 8.4 days.

In Rees [15] paper a systematic review of early evidence on LoS of patients was conducted. This paper estimates the parameters from all published papers up to 12 April 2020. According to this the mean length of stay, for countries outside of China, is 11.0 days in hospital (non it ICU) and 12.2 in ICU. The LoS in hospitalisation, and the decrease in length of stay with time in the beginning of the pandemic reported in this study is also visible in our data.

One of the limitations of the data used is the lack of distinction between length of stay in ICU care and the overall time in hospital care for a patient that received ICU treatment, as the LoS refers to the total time in hospital. Unfortunately, there are not very many studies presenting LoS, as Rees [15] also report, LoS is normally not the primary measure of interest in studies which report it, and most of them consider LoS in ICU as time in ICU hospitalisation, what makes comparing them with our data difficult. Another limitation comes from the fact that the database BI-MH does not include hospitalizations in private hospitals or other non-public healthcare providers. Other factor to take in consideration, as it overestimates entries in hospital care, is that hospital transfers count as new entries and therefore a patient can have multiple consecutive entries.

Our objective with this study was to elucidate the COVID-19 hospitalization dynamics in Portugal. We took advantage of the DRG data to explore time in hospital care and other important parameters providing the necessary tools and knowledge for the construction of better COVID-19 models that consider hospitalization. The parameters here presented were already used in the COVID-19 in-CTRL model [2].

## Data Availability

All data produced in the present work are contained in the manuscript

## ACKNOWLEDGMENTS

The authors acknowledge financial support from the Fundação para a Ciência e Tecnologia—FCT through project 692 2^<u>a</u>^ edição Research 4 covid, project name Projeção do Impacte das medidas Não-farmacológicas de Controlo e mitigação da epidemia de COVID-19 em Tempo ReaL (COVID-19 in-CTRL). The first author also acknowledges FCT within the grant with reference “COVID inCTRL/DEP/01/2020”. The second author also acknowledges FCT within the PhD grants “DOCTOR-ATES 4 COVID-19”, number 2020.10172.BD. The third author also acknowledges FCT within the PhD grant “DOCTORATES 4 COVID-19” with reference 2020.10252.BD. The fourth also acknowledges FCT within the Strategic Project UIDB/00297/2020 (Centro de Matemática e Aplicações, FCT NOVA). The fifth author also acknowledges FCT within projects UIDB/04621/2020 and UIDP/04621/2020.

### A. APPENDIX

#### A.1. Data processing

The original Portuguese DRG data used in this was extracted from the BIMH platform of the SPMS/ACSS services of Portugal and provided to us with the following structure:

The original DRG data, explained in the previous table 1, was transformed before use as to have the variable identified in table 2. The next list presents all modifications that were performed:

- From the “Health institution” column, all institutions were distributed by the ARS (Regional Health Administration) to which they belong and grouped in the “Region” column.
- From the “Date of birth” column the age of the patient at the time of entry was determined and introduced in the “Age” column;
- The “ICU” column had the name changed to “Local” and the designations changed, with “non-ICU” representing non-ICU patients and “ICU” for patients that received any ICU care;
- A column with the length of stay was added, being calculated by subtracting the entry date to the discharge date;
- Using the “Age” column, the patients where divided by age groups in a column named “Group”, with ages from 0 to 80 housed in 5 year gaps groups and the 61 and up being grouped in a single group;
- Unused columns were removed in order to facilitate the analysis.

#### A.2. Variables identification

In table 2 the variables used in the study are explained. They where obtained from the restructuring of the original DRG data, as is explained in section A.1.

The data obtained was grouped by gender, age group, time and region, for either non-ICU and ICU patients, having obtained the table 3, with the entries variable corresponding to the admission date and fatalities using the discharge date.

The regions represent the ARS to which the hospital where the entry occurred belongs to. With the population numbers used being relative to the population that is covered by each region (link1). Data from the Portuguese autonomous regions where not considered.

The total number of population by gender refers to the number of residents, for Continental Portugal, according to the Portuguese 2021 census data. With the total population being 9,857,593 people (link3). The number of individuals for each age group corresponds to the annual mean resident population by age group, according to the provisional resident population indicator, obtained from the 2021 census. The sum of the population by group is 10 298 252, being slightly lower that the, more up to date, total population relative to the 2021 preliminary census data.

The Fatalities corresponds to the date of discharge for patients that died. With the fatality rate being calculated as:

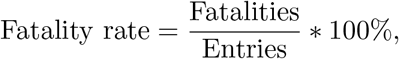

and the percentage of ICU patients as:

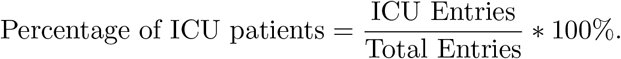

#### A.3. Representation problem

As each entry is only introduced in the database when the patient receives hospital discharged, there is a need to cut some days from the end of the data timeline, to ensure a correct representation of the data. In order to do that we calculated the mean, median and the quantiles for the length of stay, before trimming the data, for either hospitalisation and ICU (table 9).

Therefore if we remove the last 63 days from the data, we ensure that at least 95% of patients in hospital at any day are present in the data. To facilitate the analysis, instead of removing the last 63 days, we only consider entries with date of entry until 31 of March 2021. Due to the low numbers of entries, dates before 1 of March 2020 were removed. The first case of COVID-19 in Portugal was confirmed by the Portuguese authorities at March 2nd 2020, but a couple of entries with date of entry prior to that date appear in the data, this could be explained by in hospital infections. As a consequence from the original 55827, 1142 entries were removed, being considered a total of 54685 entries.

#### A.4. Summary statistics

The first summary statistics (table 10) related to the length of stay, for either type of hospitalisation was used in figure 1 to plot the violin/box plot and to expands the information visible in the histograms as they are cut at 100 days of length of stay.

Table 11 shows the summary statistics of the length of stay for patients that died, corresponding to the time until death (TuD), and for patients that where discharged from hospital, corresponding to the Time until Discharge (TuDisc). These statistics are complementary to the figure 2 violin/box plot, where a difference in LoS between groups with different outcome from hospitalisation seems to arise.

Tables with the summary statistics for LoS (table 12) and TuD (table 13), in respect to the month (month of entry for LoS and of exit for TuD), age group, gender and region, with violin/box plots build from LoS and TuD in figure 14.

#### A.5. Additional plots

#### A.6. Distribution fitting

#### A.7. Links

In the main text of the paper links for websites are provided, them being:

- **link1:** https://www.acss.min-saude.pt/wp-content/uploads/2016/10/SNS-RH-ARS-n1-dados-a-junho-de-2016.pdf;
- **link2:** https://www.ine.pt/xportal/xmain?xpid=INE&xpgid=ine_indicadores&indOcorrCod=0011166&xlang=pt;
- **link3:** https://www.ine.pt/xportal/xmain?xpid=INE&xpgid=ine_indicadores&contecto=pi&indOcorrCod=0011166&selTab=tab0.

